# Core outcome sets for trials of interventions to prevent and to treat multimorbidity in low- and middle-income countries: the COSMOS study

**DOI:** 10.1101/2024.01.29.24301589

**Authors:** Aishwarya Lakshmi Vidyasagaran, Rubab Ayesha, Jan Boehnke, Jamie Kirkham, Louise Rose, John Hurst, J. Jaime Miranda, Rusham Zahra Rana, Rajesh Vedanthan, Mehreen Faisal, Najma Siddiqi, The COSMOS Collaboration

## Abstract

**Introduction:** The burden of multimorbidity is recognised increasingly in low- and middle-income countries (LMICs), creating a strong emphasis on the need for effective evidence-based interventions. A core outcome set (COS) appropriate for the study of multimorbidity in LMIC contexts does not presently exist. This is required to standardise reporting and contribute to a consistent and cohesive evidence-base to inform policy and practice. We describe the development of two COS for intervention trials aimed at the prevention and treatment of multimorbidity in LMICs.

**Methods:** To generate a comprehensive list of relevant prevention and treatment outcomes, we conducted a systematic review and qualitative interviews with people with multimorbidity and their caregivers living in LMICs. We then used a modified two-round Delphi process to identify outcomes most important to four stakeholder groups with representation from 33 countries (people with multimorbidity/caregivers, multimorbidity researchers, healthcare professionals, and policy makers). Consensus meetings were used to reach agreement on the two final COS. Registration: https://www.comet-initiative.org/Studies/Details/1580.

**Results:** The systematic review and qualitative interviews identified 24 outcomes for prevention and 49 for treatment of multimorbidity. An additional 12 prevention, and six treatment outcomes were added from Delphi round one. Delphi round two surveys were completed by 95 of 132 round one participants (72.0%) for prevention and 95 of 133 (71.4%) participants for treatment outcomes. Consensus meetings agreed four outcomes for the prevention COS: (1) Adverse events, (2) Development of new comorbidity, (3) Health risk behaviour, and (4) Quality of life; and four for the treatment COS: (1) Adherence to treatment, (2) Adverse events, (3) Out-of-pocket expenditure, and (4) Quality of life.

**Conclusion:** Following established guidelines, we developed two COS for trials of interventions for multimorbidity prevention and treatment, specific to LMIC contexts. We recommend their inclusion in future trials to meaningfully advance the field of multimorbidity research in LMICs.

**KEY MESSAGES:** What is already known on this topic?

Although a Core Outcome Set (COS) for the study of multimorbidity has been previously developed, it does not include contributions from low- and middle-income countries (LMICs). Given the important differences in disease patterns and healthcare systems between high-income country (HIC) and LMIC contexts, a fit-for-purpose COS for the study of multimorbidity specific to LMICs is urgently needed.

**What this study adds:**

Following rigorous guidelines and best practice recommendations for developing COS, we have identified four core outcomes for including in trials of interventions for the prevention and four for the treatment of multimorbidity in LMIC settings.
The outcomes ‘Adverse events’ and ‘Quality of life (including Health-related quality of life)’ featured in both prevention and treatment COS. In addition, the prevention COS included ‘Development of new comorbidity’ and ‘Health risk behaviour’, whereas the treatment COS included ‘Adherence to treatment’ and ‘Out-of-pocket expenditure’ outcomes.

**How this study might affect research, practice, or policy:**

The multimorbidity prevention and treatment COS will inform future trials and intervention study designs by helping promote consistency in outcome selection and reporting.
COS for multimorbidity interventions that are context-sensitive will likely contribute to reduced research waste, harmonise outcomes to be measured across trials, and advance the field of multimorbidity research in LMIC settings to enhance health outcomes for those living with multimorbidity.

## INTRODUCTION

Multimorbidity, defined as living with two or more long-term health conditions (1–3), is a growing public health challenge across the world (4–6). In low- and middle-income countries (LMICs), the pooled prevalence of multimorbidity in community settings is estimated to be around 30% (7). It is associated with considerable financial burden (8) and healthcare utilisation that creates strain on often poorly-resourced health systems (9). In addition, multimorbidity occurs at younger ages in LMICs, reducing quality of life, productivity, and life expectancy (10).

To prevent and improve the treatment of multimorbidity, evidence-based interventions are needed. However, the current heterogeneity of outcomes reported in trials and uncertainty about what should be measured hamper research efforts and limit the ability to compare and synthesise evidence of effectiveness across studies and settings (11). Also, the choice of outcomes tends to be driven by researchers’ interests, leading to concerns that the measured outcomes are more important to certain stakeholders, notably researchers and health professionals, people with lived experience of multimorbidity (12). This may be particularly the case in LMICs, where the patient and carer voice in health research and representation in research processes are often limited (13, 14), or can be marginalised due to challenges such as limited health literacy, low socioeconomic status, cultural stigma, and uncertain roles (15).

Core Outcome Sets (COS) are a minimum set of outcomes (i.e., measurements or observations used to capture the effect of interventions (16)) agreed by a range of stakeholders to be the most important for measuring and reporting in *all* studies relating to a specific health condition (17). The Core Outcome Measures in Effectiveness Trials (COMET) Initiative has developed rigorous methods for COS identification that are continuously updated (18, 19). For studies addressing multimorbidity, a COS has been previously developed (20). However, it focused only on treatment and did not include prevention outcomes. Importantly, its preparatory work to identify candidate outcomes drew on published research mainly from North America (21). Further, the Delphi panel used to achieve consensus on the final COS did not have representation from LMIC contexts. These gaps are important to address, given that both health and economic data pertaining to multimorbidity suggest that prevention may be the best course of action (22). In addition, there are marked differences between high-income countries (HIC) and LMIC contexts in populations, healthcare systems, resources, the prevalence and presentation of health conditions, and the roles of family members and caregivers (23, 24). Outcomes identified as important in HICs may not be as relevant in LMIC contexts. Therefore, we aimed to develop two COS for future intervention studies relating to i) prevention and ii) treatment of multimorbidity among adults residing in LMICs.

## METHODS

We followed best practices for COS development, as set out in the COMET guidelines (16). We report our steps using the Core Outcome Set-STandards for Reporting (COS-STAR) statement (25) (Appendix 1). The COSMOS project is registered with the COMET Initiative (https://www.comet-initiative.org/Studies/Details/1580).

Our COS development involved two main stages: (i) outcome generation stage (identifying a long-list of potential outcomes that have been or could be measured in trials) through systematic review and qualitative interviews, followed by (ii) an agreement stage on the relative importance of identified outcomes for inclusion in the COS, through Delphi surveys and consensus meetings. Outcomes relevant to the prevention and treatment of multimorbidity were considered separately at each of the stages. The overall study was guided by an expert group, which included global health multimorbidity researchers, clinicians, experts in COS development methods, as well as people from LMICs with lived experience of multimorbidity and carer representatives. The main steps of the different stages are described below, and the published protocol provides further details (26).

### Outcome generation stage

#### Systematic review

We conducted a systematic review with a pre-registered protocol (PROSPERO: CRD42020197293) to identify outcomes reported in published trials and trial registrations of interventions for the prevention and treatment of multimorbidity in LMICs. Randomised (individual, cluster, and cross-over) studies of interventions (pharmacological, non-pharmacological, simple, and complex) for multimorbidity in adults (≥18 years) at risk of, or living with multimorbidity, in community, primary care, and hospital settings in LMICs were eligible for inclusion.

The search strategy was developed by an information specialist (JW) with inputs from research experts on multimorbidity in LMICs. It included terms for multimorbidity, trial design, and terms and names of LMICs, defined according to the 2019 World Bank classification (27). We searched 15 electronic databases, including trial registries, and LMIC-specific databases, from 1990 to July 2020 (Appendix 2). Each record was independently screened by two researchers, first by title and abstract, then by full texts of potentially relevant studies. Any discordance was resolved by discussion or consultation with a third researcher when required. Data on study characteristics, outcomes, and outcome measures were extracted from included studies by one researcher, with 10% of extractions cross-verified by a senior researcher. The objective of the review was to compile a list of previously studied outcomes rather than to summarise intervention effect; therefore study quality was not assessed (26).

Separate outcome lists were generated for prevention and treatment of multimorbidity, and outcomes were removed or combined based on the following criteria: duplicates, disease-specific (rather than relevant to multimorbidity), or outcome measurement metrics/tools rather than an outcome itself (e.g., biochemical measures such as lipid profile, HbA1c, etc., and questionnaires such as Short Form Health Survey (SF-36, SF-12, etc.)).

#### Qualitative interviews

To identify outcomes of importance to people with lived experience, qualitative interviews were conducted by enrolling consenting individuals (≥18 years), either living with or caring for someone with multimorbidity. Participants were selected from across a range of LMICs in diverse geographic locations. We used our existing research networks and partnerships to identify in-country research teams with experience of conducting interviews and available to perform data collection. Eligible participants were purposely recruited by these teams to achieve optimal variation according to age (over/under 65 years), sex (male/female), and type of healthcare utilisation (community or primary care/secondary or specialist care).

An information sheet written in plain language was provided to all participants to clarify concepts of outcomes and COS. Informed consent (written or recorded) in the local language was obtained prior to conducting interviews. A semi-structured interview guide was used, which was developed in English and translated into the appropriate local languages using standard forward and back translation techniques. The main topics included participants’ experience of living with [or caring for someone living with] multimorbidity and their view on what matters as the result of interventions to prevent or treat and/or care for their conditions. The interview schedule was published as part of the protocol (26). Interviews were conducted in-person by qualified interviewers in local languages and either audio-recorded or if not possible (because of technology limitations or the participant withholding consent), recorded in detailed interviewer notes. Sections of the recordings pertaining to health outcomes were transcribed manually by the local teams and translated into English. Anonymised transcripts were sent to the COSMOS team in York for analysis. Three team members (HK, JRB, RA) reviewed the extracted statements and identified individual multimorbidity prevention and treatment outcomes from them following iterative discussion.

Outcomes identified by the systematic review and interviews were assigned to either prevention or treatment lists or both as appropriate. Lay descriptions were constructed for each outcome and reviewed before finalisation to ensure understanding across all stakeholder groups. Lastly, the prevention and treatment outcomes were categorised for presentation to the Delphi panels, using Dodd’s outcome taxonomy comprising 38 categories across five core areas, namely death, physiological/clinical, life impact, resource use, and adverse events (28).

### Agreement stage

#### Delphi surveys

We conducted two rounds of online Delphi surveys to reach consensus on the importance of each outcome identified by the outcome generation stage; separate surveys were conducted for prevention and treatment outcomes. Participants were purposively sought from across four stakeholder groups, namely (i) people living with multimorbidity and their caregivers, (ii) healthcare professionals, (iii) policy makers, and (iv) multimorbidity researchers. The identification and recruitment of participants used multiple strategies such as broadcasting through a project Twitter account, patient and public involvement groups, and COSMOS team networks (including other global health research groups, professional societies, non-government organisations relevant to multimorbidity, and government ministries). Additional strategies to recruit healthcare professionals and multimorbidity researchers included personalised emails sent to corresponding authors of studies included in our systematic review and flyers posted in partner research organisations.

We used the DelphiManager 5.0 platform, developed and maintained by the COMET Initiative (University of Liverpool) (18), to administer all surveys. The order of presenting outcome domains (based on Dodd’s taxonomy) was randomised to reduce bias. A lay description for each outcome was provided. Survey participants were asked to score the importance of each outcome for inclusion in the prevention and treatment COS, without considering its feasibility or measurability. For scoring, the GRADE (Grading of Recommendations Assessment, Development and Evaluations) 9-point Likert scale was used, with the following categories (16): ‘Not Important’ (scores 1-3), ‘Important but Not Critical’ (4–6), and ‘Critical for Inclusion’ (7–9). There was also an ‘Unable to Score’ response option, as well as the opportunity to suggest additional outcomes. For Delphi round one, for each outcome, we determined the proportion of scores in each of the GRADE categories, both overall and for each stakeholder group. All additional outcomes suggested by survey participants were reviewed for duplication and relevance by the research team, and those eligible (new distinct outcomes relevant to multimorbidity studies) were included in the Delphi round two surveys.

For Delphi round two, participants received their own round one scores, as well as the summary scores (overall and for each stakeholder group), with visual representation using histograms of the proportion of scores in each GRADE category. Participants were asked to re-score the importance of each outcome using the same 9-point Likert scale and to provide free-text reasons for any changes. Reminder emails were sent for both Delphi rounds until a minimum acceptable level of participation was achieved (70% overall, as advised by COS experts). Ratings from round two were analysed and summarised as for round one under the GRADE categories. Outcomes were then grouped according to the consensus definitions recommended by COMET (see Table 1) (11). To aid understanding of the findings and indicate clearly where there was consensus across stakeholder groups (or its absence), outcomes were presented in colour-coded tables (Table 1).

**Table 1.** Criteria for categorisation of outcomes in the Delphi surveys.

|  |  |
| --- | --- |
| Green | Outcomes scored 'Critical for Inclusion' (7-9) by $\geq 70\%$ of respondents in all 4 stakeholder groups. |
| Purple | Outcomes scored 'Critical for Inclusion' (7-9) by $\geq 70\%$ of respondents in 3 of 4 stakeholder groups. |
| Blue | Outcomes scored 'Critical for Inclusion' (7-9) by $\geq 70\%$ of respondents in 2 of 4 stakeholder groups. |
| Yellow | Outcomes scored 'Critical for Inclusion' (7-9) by $\geq 70\%$ of respondents in 1 of 4 stakeholder group. |
| Red | Outcomes scored 'Critical for Inclusion' (7-9) by $< 70\%$ of the respondents in all 4 stakeholder groups. |

#### Consensus meetings

All Delphi participants were sent electronic invitations for the consensus meetings. A modified nominal group technique was used to discuss findings from the Delphi surveys and to develop agreements on critical outcomes for inclusion in the COS (16, 29, 30). Separate meetings were held for prevention and treatment outcomes, using the Zoom online platform (31) to maximise participation from multiple countries. Two pre-meeting sessions were held to orientate attendees to the purpose of the consensus meetings, scope of the COS, and the use of Zoom. In addition, an information pack describing the processes followed in the study and presenting results from the Delphi surveys using colour-coded tables (as described above), were sent to all participants before the meetings. Those participants unable to attend the virtual meetings were invited to send in their views by email.

At the start of each meeting, we reminded participants of the aim (i.e., developing consensus on the inclusion of outcomes in the COS), and outlined the meeting structure and process to ensure inclusive discussions. Meetings were facilitated by experts with extensive experience in COS development (JK, LR). Results from the Delphi surveys were presented. Outcomes scored as ‘Critical for Inclusion’ by ≥70% of Delphi respondents in all 4 stakeholder groups (colour-coded green, see Table 1) were included in the COS if they met the consensus meeting threshold of ≥80% voting for inclusion; otherwise, they underwent further discussion. Outcomes scored as ‘Critical for Inclusion’ by ≥70% of Delphi respondents in only one or no stakeholder groups (colour-coded yellow/red) were excluded without further discussion unless nominated to be ‘saved’ and supported by voting above a threshold of

≥80% by meeting participants. All outcomes scored as ‘Critical for Inclusion’ by ≥70% of Delphi respondents in two or three stakeholder groups (colour-coded blue/purple), were discussed further. Views shared by email by individuals unable to attend meetings were also fed into the meeting. Iterative rounds of whole-group and small-group discussions, facilitated by Google Jamboard, were used to categorise the outcomes for discussion into ‘Critical’ ‘Good to include’ and Not Important’. Discussions were followed by voting to include or exclude outcomes in the COS.

Following the consensus meetings, participants were emailed for a further vote on any outcomes for which consensus was not reached during the meetings, and for feedback on the wording and descriptions of outcomes voted for inclusion. The two final COS for prevention and treatment were compiled and sent to all consensus meeting participants for final endorsement.

### Ethics and permission for data collection

Ethics approval for the research was obtained from the Health Sciences Research Governance Committee at the University of York (HSRGC/2020/409/D: COSMOS). Approvals were also obtained from relevant in-country ethics committees for all participating interview sites (Appendix 3). Informed consent (written or audio-recorded) in the local language was obtained from all participants prior to conducting interviews. All Delphi panellists and consensus meeting attendees also provided consent before participation.

### Patient and public involvement

Four members of the steering committee overseeing the study were people living with multimorbidity and their caregivers. The study also benefited from advice from the NIHR IMPACT in South Asia Group (https://www.impactsouthasia.com/impact-group/) Community Advisory Panels and from the NCD Alliance (https://ncdalliance.org/), a civil society network, advocating for people with non-communicable diseases.

## RESULTS

### Outcome generation stage

Figures 1a & 1b show the steps of outcome generation related to the prevention and treatment of multimorbidity, respectively.

**Figure 1a.**
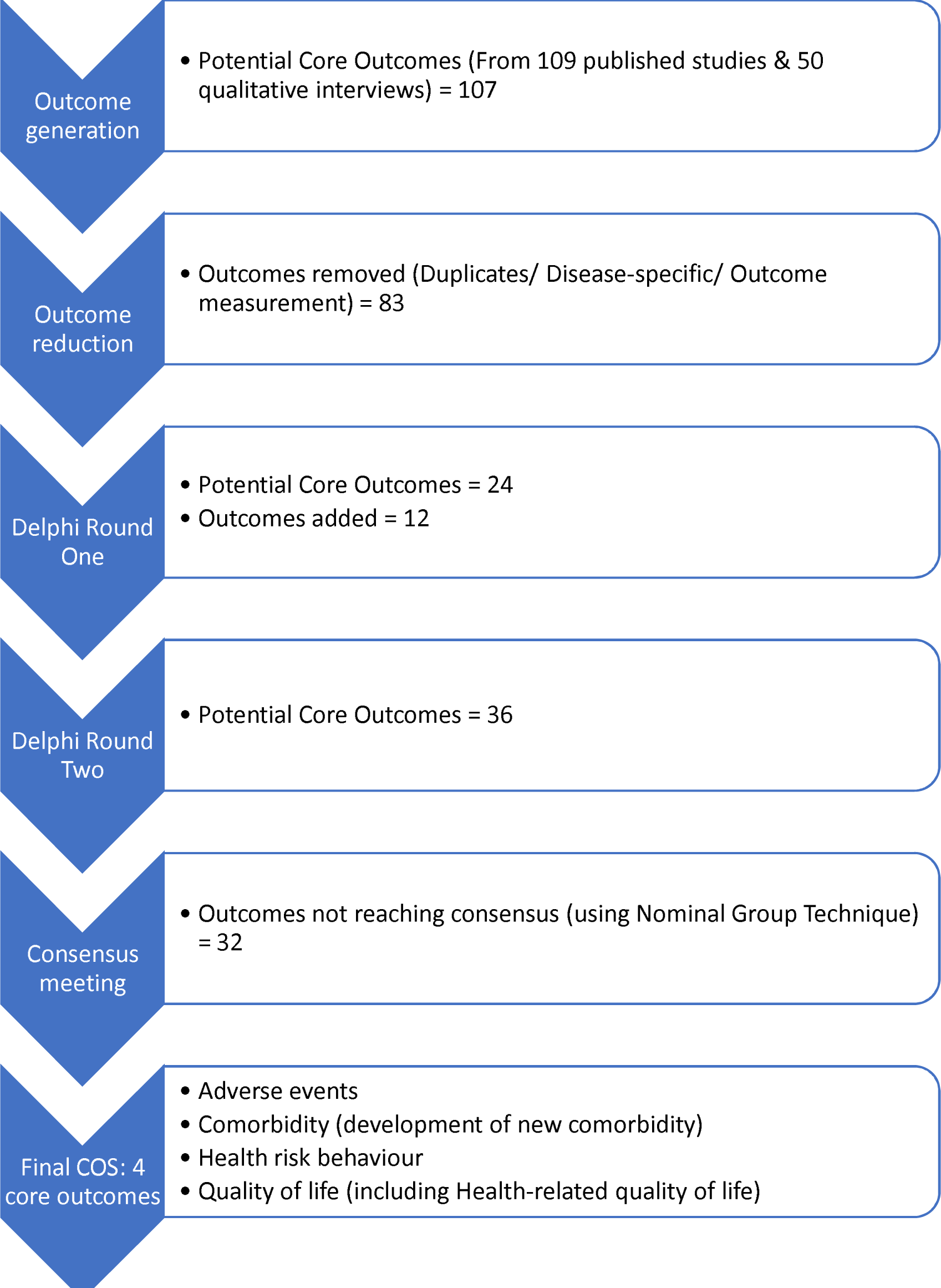
Development of COS for trials of interventions to prevent multimorbidity in LMICs.

**Figure 1b.**
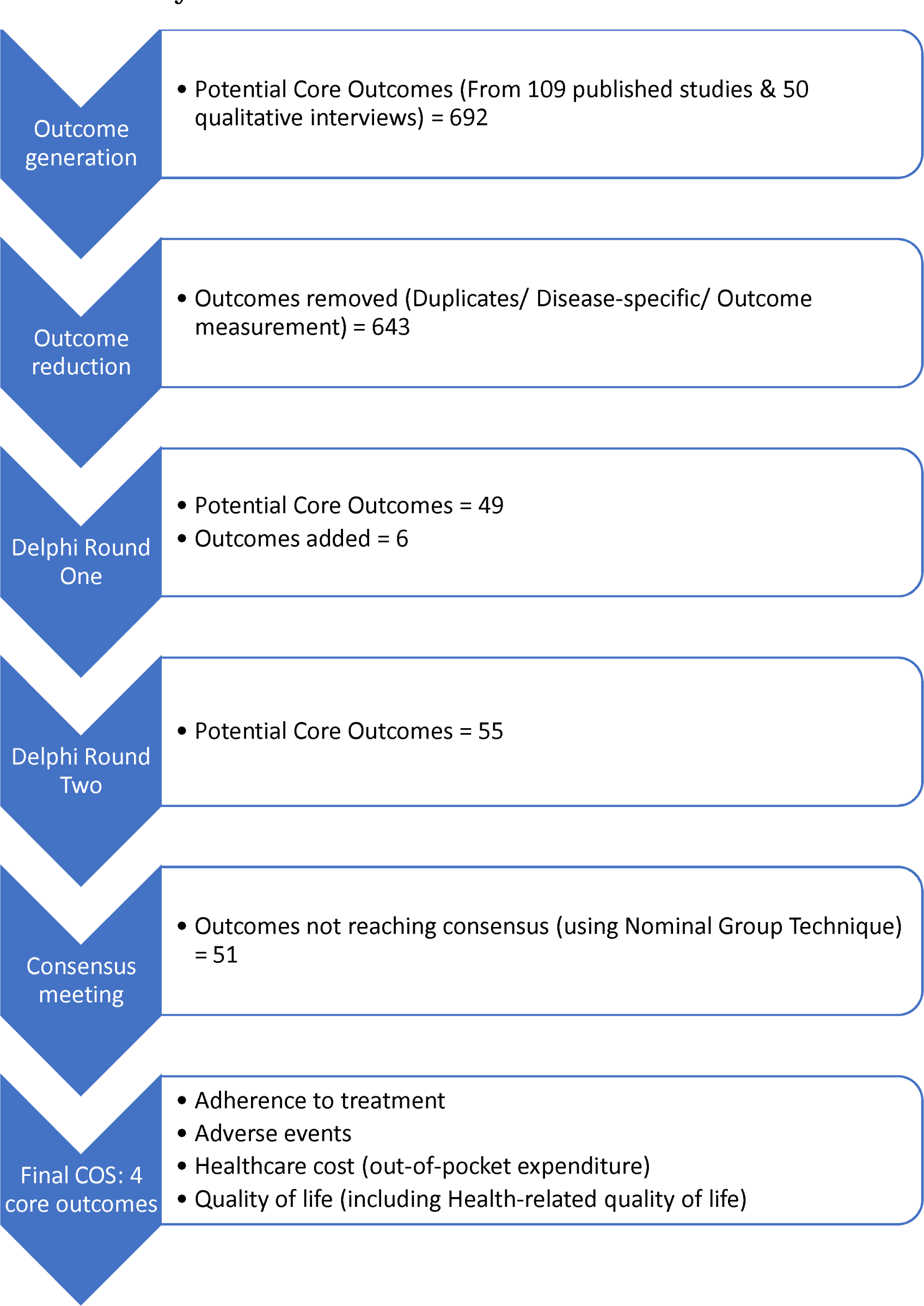
Development of COS for trials of interventions to treat multimorbidity in LMICs.

#### Systematic review

Our searches yielded 17,267 records, with 16,949 remaining after removing duplication publications (Appendix 2). Following title and abstract screening, 16,705 records were excluded, and the remaining 243 papers were obtained. Full-text screening resulted in the exclusion of a further 134 records (Appendix 2). The remaining 109 randomised intervention studies on the prevention and treatment of multimorbidity conducted in at least 25 LMICs were included.

From these papers, 92 prevention and 236 treatment outcomes were extracted and reduced to 19 prevention and 38 treatment outcomes after removing duplicate outcomes, disease-specific outcomes, and outcome measurement metrics.

#### Qualitative interviews

The interviewees included five participants from each of the following ten countries: Afghanistan and Burkina Faso (low-income), Bangladesh, Ghana, Nepal, Nigeria, and Pakistan (lower-middle income), and Mexico, Peru, and Suriname (upper-middle income), totalling 50 interviewees. They comprised 37 people living with multimorbidity and 13 family caregivers. The distribution of socio-demographic characteristics was as follows: sex (46% male and 54% female), age (80% under 65 and 20% 65+ years), and type of healthcare utilisation (34% community/primary care and 66% secondary/specialist care). Participants reported having from two to five co-existing conditions, including tuberculosis, asthma, hypertension, diabetes, cardiovascular disease, HIV, cancer, stroke, COPD, mental health disorders and others.

The interviews generated five further outcomes for prevention and 11 for treatment of multimorbidity (see Appendix 4 for example coding of qualitative data). Combining these outcome lists with the corresponding lists generated from the systematic review resulted in 24 outcomes for prevention and 49 for treatment of multimorbidity, which were classified according to Dodd’s taxonomy and presented in the Delphi round one surveys (Appendix 5).

### Agreement stage

Table 2 summarises the characteristics of participants in the Delphi surveys and consensus meetings; Table 3 shows the outcomes scored as critical for inclusion at each of the agreement stages.

**Table 2.** Characteristics of participants in Delphi surveys and consensus meetings.

| Name of survey/ meeting | Stakeholder group, n (%) | Age group, n (%) | Female, n (%) | Region, n (%) |
| --- | --- | --- | --- | --- |
| Delphi round 1, prevention survey (N = 132) | People living with multimorbidity/Caregivers = 29 (22.0)<br>Healthcare professionals = 27 (20.4)<br>Policy makers = 12 (9.1)<br>Multimorbidity researchers = 64 (48.5) | 18-24 = 4 (3.0)<br>25-34 = 34 (25.7)<br>35-44 = 34 (25.7)<br>45-54 = 26 (19.7)<br>55-64 = 24 (18.2)<br>65+ = 10 (7.6) | 68 (51.5) | Low income = 6 (4.5)<br>Lower middle income = 74 (56.1)<br>Upper middle income = 15 (11.4)<br>High income = 37 (28.0) |
| Delphi round 1, treatment survey (N = 133) | People living with multimorbidity/Caregivers = 30 (22.5)<br>Healthcare professionals = 24 (18.0)<br>Policy makers = 13 (9.8)<br>Multimorbidity researchers = 66 (49.6) | 18-24 = 3 (2.2)<br>25-34 = 36 (27.0)<br>35-44 = 35 (26.3)<br>45-54 = 26 (19.5)<br>55-64 = 22 (16.5)<br>65+ = 11 (8.4) | 70 (52.6) | Low income = 4 (3.0)<br>Lower middle income = 78 (58.6)<br>Upper middle income = 17 (12.8)<br>High income = 34 (25.6) |
| Delphi round 2, prevention survey (N = 95) | People living with multimorbidity/Caregivers = 24 (25.3)<br>Healthcare professionals = 14 (14.7)<br>Policy makers = 8 (8.4)<br>Multimorbidity researchers = 49 (51.6) | NA | 49 (51.6) | Low income = 5 (5.3)<br>Lower middle income = 45 (47.4)<br>Upper middle income = 10 (10.5)<br>High income = 35 (36.8) |
| Delphi round 2, treatment survey (N = 95) | People living with multimorbidity/Caregivers = 22 (23.1)<br>Healthcare professionals = 16 (16.8)<br>Policy makers = 10 (10.5)<br>Multimorbidity researchers = 47 (49.5) | NA | 51 (53.7) | Low income = 5 (5.3)<br>Lower middle income = 48 (50.5)<br>Upper middle income = 11 (11.6)<br>High income = 31 (32.6) |
| Consensus meeting, prevention (N = 19) | People living with multimorbidity/Caregivers = 3 (15.8)<br>Healthcare professionals = 4 (21.0)<br>Policy makers = 1 (5.3)<br>Multimorbidity researchers = 11 (57.9) | NA | 10 (52.6) | Low income = 1 (5.3)<br>Lower middle income = 11 (57.9)<br>Upper middle income = 1 (5.3)<br>High income = 6 (31.6) |
| Consensus meeting, treatment (N = 14) | People living with multimorbidity/Caregivers = 1 (7.1)<br>Healthcare professionals = 4 (28.6)<br>Policy makers = 1 (7.1)<br>Multimorbidity researchers = 8 (57.1) | NA | 7 (50.0%) | Low income = 0 (0.0)<br>Lower middle income = 6 (42.8)<br>Upper middle income = 4 (28.6)<br>High income = 4 (28.6) |

**Table 3.** Critical outcomes selected in Delphi surveys and consensus meetings.

| Delphi round 1 | Delphi round 2 | Delphi results | Consensus |
| --- | --- | --- | --- |
| **Prevention outcomes** |  |  |  |
| **‘Critical for Inclusion’ by ≥70% of all participants** | **Voted ‘Critical for Inclusion’ (7-9) by ≥70% of all participants** | **‘Critical for Inclusion’ by ≥70% in 4 (green), 3 (purple), and 2 (blue) stakeholder groups** | **Prevention COS** |
| 1. Adherence to treatment<br>2. Adverse events<br>3. Cardiovascular event<br>4. Cardiovascular risk<br>5. Cognitive function<br>6. Comorbidity<br>7. Cost effectiveness<br>8. Death<br>9. Health-related quality of life<br>10. Obesity<br>11. Organ damage<br>12. Prevention of hypertension<br>13. Psychological wellbeing<br>14. Quality of life<br>15. Timely screening | 1. Adherence to treatment<br>2. Adverse events<br>3. Cardiovascular event<br>4. Cardiovascular risk<br>5. Chronic disease self-management*<br>6. Comorbidity<br>7. Cost effectiveness<br>8. Death<br>9. Health risk behaviour*<br>10. Health-related quality of life<br>11. Obesity<br>12. Organ damage<br>13. Prevention of hypertension<br>14. Psychological wellbeing<br>15. Quality of life<br>16. Timely screening | <u>Green-coded outcomes:</u><br>1. Adverse events<br>2. Cardiovascular event<br>3. Chronic disease self-management*<br>4. Comorbidity<br>5. Prevention of hypertension<br>6. Quality of life<br><u>Purple-coded outcomes:</u><br>1. Cost effectiveness<br>2. Death<br>3. Obesity<br>4. Organ damage<br>5. Pain<br>6. Psychological wellbeing<br>7. Timely screening<br><u>Blue-coded outcomes:</u><br>1. Adherence to treatment<br>2. Cardiovascular risk<br>3. Cognitive function<br>4. Diet<br>5. Early detection*<br>6. Health risk behaviour*<br>7. Health-related quality of life<br>8. Treatment satisfaction | 1. Adverse events<br>2. Comorbidity (development of new comorbidity)<br>3. Health risk behaviour*<br>4. Quality of life (including Health-related quality of life) |
| **Other outcomes** | **Other outcomes** | **‘Critical for Inclusion’ by ≥70% in 1 (yellow), or none (red) of the stakeholder groups** | **Not in COS, but suggested as additional outcome** |
| 1. Diet<br>2. Exercise tolerance<br>3. Fatigue<br>4. Health literacy<br>5. Healthcare use | 1. Carer burden*<br>2. Cognitive function<br>3. Diet<br>4. Early detection*<br>5. Exercise tolerance | <u>Yellow-coded outcomes:</u><br>1. Carer burden*<br>2. Health literacy<br>3. Self-efficacy*<br>4. Weight | 1. Healthcare use (including cost effectiveness) |
| 6. Pain<br>7. Reduced medication<br>8. Treatment satisfaction<br>9. Weight | 6. Fatigue<br>7. Functioning/ADL*<br>8. Health anxiety*<br>9. Health literacy<br>10. Health seeking behaviour*<br>11. Healthcare use<br>12. Income*<br>13. Loneliness*<br>14. Pain<br>15. Perceived health*<br>16. Reduced medication<br>17. Self-efficacy*<br>18. Social functionality*<br>19. Treatment satisfaction<br>20. Weight | <u>Red-coded outcomes:</u><br>1. Exercise tolerance<br>2. Fatigue<br>3. Functioning/ADL*<br>4. Health anxiety*<br>5. Health seeking behaviour*<br>6. Healthcare use<br>7. Income*<br>8. Loneliness*<br>9. Perceived health*<br>10. Reduced medication<br>11. Social functionality* |  |
| **Treatment outcomes** |  |  |  |
| **‘Critical for Inclusion’ by ≥70% of all participants** | **‘Critical for Inclusion’ by ≥70% of all participants** | **‘Critical for Inclusion’ by ≥70% in 4 (green), 3 (purple), and 2 (blue) stakeholder groups** | **Treatment COS** |
| 1. Adherence to treatment<br>2. Adverse events<br>3. Cardiovascular event<br>4. Cognitive function<br>5. Comorbidity<br>6. Cost effectiveness<br>7. Death<br>8. Healthcare access<br>9. Healthcare cost<br>10. Healthcare quality<br>11. Health-related quality of life<br>12. Increase in symptoms<br>13. Psychological wellbeing<br>14. Quality of life<br>15. Treatment satisfaction | 1. Adherence to treatment<br>2. Adverse events<br>3. Cardiac event risk<br>4. Cardiovascular event<br>5. Cognitive function<br>6. Comorbidity<br>7. Cost effectiveness<br>8. Death<br>9. Healthcare access<br>10. Healthcare cost<br>11. Healthcare quality<br>12. Health-related quality of life<br>13. Hospital admission<br>14. Illness under control<br>15. Psychological wellbeing<br>16. Quality of life<br>17. Treatment satisfaction | <u>Green-coded outcomes:</u><br>1. Adherence to treatment<br>2. Death<br>3. Healthcare access<br>4. Healthcare cost<br>5. Healthcare quality<br><u>Purple-coded outcomes:</u><br>1. Adverse events<br>2. Cardiovascular event<br>3. Comorbidity<br>4. Cost effectiveness<br>5. Health-related quality of life<br>6. Healthcare staff communication<br>7. Increase in symptoms<br>8. Pain<br>9. Psychological wellbeing<br>10. Quality of life<br>11. Treatment burden**<br>12. Treatment satisfaction<br><u>Blue-coded outcomes:</u><br>1. Cardiovascular risk<br>2. Cognitive function<br>3. Continuity of care**<br>4. Falls risk<br>5. Health risk behaviour<br>6. Healthcare use<br>7. Hospital admission | 1. Adherence to treatment<br>2. Adverse events<br>3. Healthcare cost (out-of-pocket cost of treatment)<br>4. Quality of life (including Health-related quality of life) |
|  |  | 8. Hypertension<br>9. Illness under control<br>10. Obesity<br>11. Perceived health<br>12. Physical activity |  |
| **Other outcomes** | **Other outcomes** | **‘Critical for Inclusion’**<br>**by ≥70% in 1 (yellow),**<br>**or none (red) of the**<br>**stakeholder groups** | **Not in COS, but**<br>**suggested as additional**<br>**outcome** |
| 1. Acceptance of illness<br>2. Aggression<br>3. Agitation<br>4. Appetite<br>5. Balance<br>6. Cardiac event risk<br>7. Carer burden<br>8. Diet<br>9. Domestic violence<br>10. Emotional regulation<br>11. Falls risk<br>12. Fatigue<br>13. Functioning/ADL<br>14. Health anxiety<br>15. Health literacy<br>16. Health risk behaviour<br>17. Healthcare staff<br>communication<br>18. Healthcare use<br>19. Hospital admission<br>20. Hypertension<br>21. Illness resolution<br>22. Illness stigma<br>23. Illness under control<br>24. Income<br>25. Loneliness<br>26. Nausea<br>27. Obesity<br>28. Pain<br>29. Perceived health<br>30. Physical activity<br>31. Reduced medication<br>32. Self-management<br>33. Sleep quality<br>34. Weight | 1. Acceptance of illness<br>2. Aggression<br>3. Agitation<br>4. Appetite<br>5. Balance<br>6. Carer burden<br>7. Continuity of care**<br>8. Diet<br>9. Domestic violence<br>10. Emotional regulation<br>11. Falls risk<br>12. Fatigue<br>13. Frailty**<br>14. Functioning/ADL<br>15. Health anxiety<br>16. Health literacy<br>17. Health risk behaviour<br>18. Healthcare staff<br>communication<br>19. Healthcare use<br>20. Hypertension<br>21. Illness resolution<br>22. Illness stigma<br>23. Income<br>24. Increase in symptoms<br>25. Loneliness<br>26. Nausea<br>27. Obesity<br>28. Pain<br>29. Perceived health<br>30. Physical activity<br>31. Polypharmacy**<br>32. Reduced medication<br>33. Self-esteem**<br>34. Self-management<br>35. Sleep quality<br>36. Social<br>functionality**<br>37. Treatment burden**<br>38. Weight | <u>Yellow-coded outcomes:</u><br>1. Functioning/ADL<br>2. Illness resolution<br>3. Income<br>4. Polypharmacy**<br>5. Self-management<br>6. Sleep quality<br>7. Social<br>functionality**<br><u>Red-coded outcomes:</u><br>1. Acceptance of illness<br>2. Aggression<br>3. Agitation<br>4. Appetite<br>5. Balance<br>6. Carer burden<br>7. Diet<br>8. Domestic violence<br>9. Emotional regulation<br>10. Fatigue<br>11. Frailty**<br>12. Health anxiety<br>13. Health literacy<br>14. Illness stigma<br>15. Loneliness<br>16. Nausea<br>17. Reduced medication<br>18. Self-esteem**<br>19. Weight | N/A |

\*Additional prevention outcomes suggested during Delphi round 1; \*\*Additional treatment outcomes suggested during Delphi round 1 (see Appendix 5 for help text)

#### Delphi surveys

The Delphi round one prevention and treatment surveys were completed by 132 and 133 participants, respectively, with 127 completing both. The distribution of stakeholders was similar in both groups, with multimorbidity researchers making up almost half the sample, followed by people living with multimorbidity/caregivers (∼22%), healthcare professionals (18-20%), and policy makers (<10%). Over half of the participants in both groups were 25-44 years old and women. The largest geographical representation in both groups was from lower middle-income countries (56-58%), followed by high income (25-28%), upper middle income (∼12%), and low-income countries (<5%). The Delphi round two surveys were completed by 95 participants, for prevention (72.0% of round one) and treatment outcomes (71.4% of round one). By stakeholder groups, round two completions were >70% of round one participants for all groups, except healthcare professionals (66.7% in prevention and treatment surveys) and policymakers (51.8% in prevention survey).

Of the outcomes presented in Delphi round one, 15 (of 24) prevention and 15 (of 49) treatment outcomes were rated as ‘Critical for Inclusion’ (scores 7-9) by ≥70% of all participants (Table 3). Thirty-eight additional outcomes were proposed for prevention with 12 of them included in Delphi round two after reviewing for duplication, and relevance to multimorbidity. For treatment, six of 24 proposed additional outcomes were taken forward to Delphi round two. Overall, 36 prevention (24 generated from the review and interviews, and 12 additional suggestions by Delphi round one respondents) and 55 treatment outcomes (49 generated and 6 additional suggestions) were presented for rating in the Delphi round two surveys.

In the Delphi round two surveys, 16 (of 36) prevention and 17 (of 55) treatment outcomes were rated as ‘Critical for Inclusion’ (scores 7-9) by ≥70% of all participants (Table 3).

Categorising by stakeholder groups, in the prevention list, six outcomes were coded green (‘Critical for Inclusion’ by >70% in all stakeholder groups), 15 were coded blue/purple (‘Critical for Inclusion’ by >70% in any three or two stakeholder groups), and 15 were coded yellow/red (‘Critical for Inclusion’ by >70% in one or none of the stakeholder groups); the treatment list included five (green), 31 (blue/purple), and 19 (yellow/red) outcomes, following the same categorisation (Table 3).

### Consensus meetings

#### Prevention

The consensus meeting for prevention had 17 in-meeting and two email participants (Table 2), including 11 (57.9%) multimorbidity researchers, three people living with multimorbidity/caregivers (15.8%), four healthcare professionals (21.0%), and one policy maker (5.3%). Following the nominal group technique discussions, 32 of the 36 outcomes were excluded; there was consensus on the four outcomes presented below for inclusion in the prevention COS (Table 3).

‘Comorbidity’ and ‘Quality of life’ (both green-coded outcomes) received 93% of votes in the consensus meeting, but both outcomes were recommended for further discussion regarding their wording. Following these discussions, for the prevention COS, it was agreed ‘Comorbidity’ referred to the prevention of development of a new illness alongside the existing health condition being examined in a trial; the wording of this outcome was therefore amended as ‘Comorbidity (development of new comorbidity)’. Similarly, ‘Quality of life’ was reworded as ‘Quality of life (including Health-related quality of life)’, to reflect the consensus that the two outcomes should be combined (with researchers free to choose the most appropriate measure for their trial).

‘Adverse events’ (green-coded) did not initially reach the voting threshold of ≥80%, with those opposing its inclusion suggesting that measuring such events would be common practice across studies. However, it was included following further discussion and consensus among meeting participants, who considered it incorporated a range of negative outcomes for research teams to decide as appropriate for the multimorbidity prevention intervention being implemented.

The outcome ‘Health risk behaviours’ (blue-coded), proposed for the prevention COS during the Delphi round one survey, was similarly included following consensus discussions. The term was understood to include the range of behaviours considered to be risk factors for chronic conditions such as tobacco use, physical inactivity, and unhealthy diet.

The outcome ‘Healthcare use’ (including cost-effectiveness) generated considerable debate. While it was considered very important, it was argued that it might not be relevant to all trials. ‘Healthcare use’ did not reach the voting threshold for inclusion in the COS for this reason, but the meeting consensus was that it should be recommended as an important outcome to consider in relevant trials.

#### Treatment

The treatment consensus meeting comprised 12 in-meeting and two email participants (Table 2), including 8 (57.1%) multimorbidity researchers, four healthcare professionals (28.6%), one person living with multimorbidity/caregiver (7.1%), and one policy maker (7.1%). There was consensus on the following four outcomes as critical for inclusion (Table 3).

‘Adherence to treatment’ and ‘Healthcare costs’ (both green-coded outcomes) received 90% and 100% of votes respectively; additional clarification was added to the latter, stipulating it was specifically ‘out-of-pocket expenditure’ that was considered critical in LMIC studies, and as such this should be specified in the COS. The outcomes ‘Adverse events’ and ‘Quality of life’ (including Health-related quality of life) (both purple-coded) were included after further discussion and consensus. While it was agreed that death should be reported as an adverse event where relevant, the outcome ‘Death’ or ‘Mortality’ did not reach the threshold for inclusion separately (60% votes).

## DISCUSSION

The COSMOS study followed rigorous participatory methods as recommended by COMET with representation from diverse geographies and stakeholder groups to develop two COS for use in future trials of interventions to prevent and to treat multimorbidity among adults living in LMICs. The two COS included four outcomes each, with ‘Adverse events’ and ‘Quality of life (including Health-related quality of life)’ featured in both sets. In addition, the prevention COS included ‘Development of new comorbidity’ and ‘Health risk behaviour’, whereas the treatment COS included ‘Adherence to treatment’ and ‘Out-of-pocket expenditure’ outcomes.

A previously developed COS for multimorbidity (COSmm) (20) with inputs from a systematic review of studies (21) and an expert panel, both solely from HICs, also included ‘Health-related quality of life’ among their highest-scoring outcomes. Our consensus panels voted to combine this outcome with the broader ‘Quality of life’ both in the prevention and treatment COS. This similarity between COSmm and our results suggests that the outcome ‘Quality of life’ may be relevant to multiple stakeholders and well-suited across diverse contexts to capture the impacts of living with multimorbidity. Nonetheless, further work on differentiating these constructs and their operationalisation will be necessary to translate this finding into actionable research and clinical practice (32, 33). The inclusion of ‘Adherence to treatment’ and ‘Healthcare costs’ are further similarities between COSmm and our treatment COS. However, while costs are only presented as a broad health systems outcome in COSmm, we specify its scope as covering out-of-pocket treatment costs to people living with multimorbidity, given that this can be an important source of catastrophic health expenditures and impoverishment in many LMICs (13, 34, 35).

‘Healthcare use’ was included in COSmm but did not reach a consensus for inclusion in our LMIC COS. This likely reflects differences across LMICs in the use of healthcare services (36). Additionally, the outcome may be severely limited in some LMICs due to lack of access to services (37). Nevertheless, it was considered an important outcome, which should be included (with or without cost-effectiveness) in some prevention studies, where appropriate. We considered it particularly important to develop a separate COS for the prevention of multimorbidity, given that the targets for prevention and treatment interventions are often different. In addition, as non-communicable diseases (with amenable risk factors) form a large proportion of the multimorbidity burden, a COS for prevention trials that will help build the evidence base is critical. With clear opportunities for implementing prevention strategies targeting risk factors (38), it is noteworthy that ‘Health risk behaviour’ has been included in our prevention COS.

Currently, most COS reflect priorities from HIC perspectives only, with very few including participants from LMICs and even fewer initiated in LMICs (13, 39, 40). Given that there are important differences in populations, disease patterns and healthcare systems between HIC and LMIC contexts (23, 24), our two COS for intervention studies to prevent and treat multimorbidity specifically in LMICs are likely to be more context-relevant, with greater applicability and adoptability in these settings (41). Another advantage of the COS will be more consistent and aligned outcome reporting in future multimorbidity trials, leading to systematic reviews that are more meaningful, as like-for-like outcomes can be combined in meta-analyses.

Having agreed COS for LMIC multimorbidity trials, further work is needed to review the evidence base and develop consensus on validated metrics or tools, which should be used to capture these outcomes. This was beyond the scope of the current study, but we have collated from our systematic review the measures and tools used to assess the six outcomes included in the two multimorbidity COS (see Appendix 6). Also, the large difference in the number of potential outcomes identified for prevention trials (N=107) and treatment trials (N=692) illustrates the need for more research on (and implementation of) of preventive interventions.

The strengths of our study include adherence to the recommended COMET guidelines (16) at both the outcome generation and agreement stages. We used a combination of rigorously conducted approaches (systematic review and qualitative interviews) to generate the initial lists of prevention and treatment outcomes, and multi-stage consensus building exercises involving a wide range of stakeholders across backgrounds, professions, and countries.

There are three key limitations to consider. First, unlike the interviews conducted in local languages, the Delphi surveys were administered in English, using the DelphiManager online platform, thereby limiting participation to individuals who could read or speak English and had a degree of confidence in using online tools. To mitigate the impact of this, support was provided by in-country research partners, but this was challenging to do consistently for Delphi round two, which led to higher than anticipated attrition. Nonetheless, the study achieved a satisfactory response rate in the round two surveys (>70.0% of round one participants for both prevention and treatment rounds), with representation from across 33 countries, which may not have been possible without using online tools.

Another limitation was that multimorbidity researchers were the largest stakeholder group in the agreement stages, with the risk that the consensus and final COS might largely reflect their views. Policymakers, on the other hand, had the least representation. However, our approach ensured that views were included from all four stakeholder groups at all consensus-building stages (Table 2). Delphi survey responses were summarised by stakeholder group, with agreement across groups being a key consideration in identifying outcomes as important. The selection of outcomes for the COS in consensus meetings also took account of their importance for all stakeholder groups.

Lastly, methods for COS development are evolving (42). While our approach adheres to the currently recommended steps and represents an advance over the previous COS for multimorbidity, the evidence base for developing consensus is limited (43) (for example, on the optimum way to present results in Delphi surveys, or to conduct discussions and achieve equitable, inclusive ranking or voting on outcomes). We further acknowledge that continued efforts are needed to understand the uptake and impact of COS, as demonstrated in other areas of health (44).

In addition, the definition of an intervention to prevent and/or treat multimorbidity might itself need further development (45). Repeatedly identified issues in the management of multimorbidity are the lack of integrated care and inadequate considerations of cross-treatment interactions, complications, and consequences (46). Interventions that consider these issues might be ones which have a planned positive impact on one or more conditions, while considerations are undertaken to minimise, reduce or avoid negative impacts from the presence of multimorbidity. Future efforts may be needed to include this broader scope.

## CONCLUSION

In conclusion, the COSMOS study has developed two COS specifically for LMICs, to include in all intervention studies focusing on the prevention and treatment of multimorbidity. The two COS comprise four outcomes each, carefully selected using recommended standards, and therefore likely to be relevant and meaningful to a wide range of LMIC stakeholders, including people living with multimorbidity, their caregivers, multimorbidity researchers, healthcare professionals, and policy makers. Future research should identify and develop consensus on validated measures to assess these outcomes.

Uptake of COS in future trials will promote consistency in outcome selection and reporting and thereby ensure the comparability of effectiveness across different studies on multimorbidity in LMICs.

## Data Availability

All data produced in the present study are available upon reasonable request to the authors

## ACKNOWLEDGEMENTS

Funding for the DelphiManager software licence was provided by the Global Alliance for Chronic Diseases (GACD). We also thank the GACD for publicising the Delphi exercises across their networks.

We thank all the patient advisers and caregivers for their participation at different stages of the study. We also thank all the researchers, healthcare professionals, and policy makers who participated in the Delphi surveys and consensus meetings.

## AUTHOR CONTRIBUTION STATEMENT

`o` Conceptualization –Jan R Boehnke, Najma Siddiqi
`o` Design of protocol – Alyssa Chase-Vilchez, Corrado Barbui, Eleonora Uphoff, Gerardo A Zavala, Gina Agarwal, Jamie J Kirkham, Jan R Boehnke, John R Hurst, Josefien van Olmen, Judy Wright, Kamran Siddiqi, Louise Rose, Marianna Purgato, Najma Siddiqi, Naomi Levitt, Oscar Flores-Flores, Rachel Churchill, Rajesh Vedanthan, Rusham Zahra Rana
`o` Conduct of systematic review – A Cristina Garcia-Ulloa, Augustine Nonso Odili, Darwin Del Castillo, Eleonora Uphoff, Gerardo Zavala Gomez, Judy Wright, Kamrun N Koly, Koralagamage Kavindu Appuhamy, Najma Siddiqi, Rubab Ayesha, Sesergio Baldew
`o` Conduct of qualitative study – A Cristina Garcia-Ulloa, Adewale L Oyeyemi, Asiful Haider Chowdhury, Cecilia Anza Ramirez, Dolores Ronquillo, Humaira Khalid, Jan R Boehnke, Johnblack Kabukye, Josefien van Olmen, Jose Patricio López-Jaramillo, Joseph Senyo Kwashie, Kingsley Akinroye, Laura Downey, María del Carmen Caamaño, Marianna Purgato, Naomi Levitt, Niels Victor Pacheco Barrios, Olga P García, Pervaiz Tufail, Phuong Tran Bich, Qirat Naz, Rakesh Singh, Rubab Ayesha, Rufus Olusola Akinyemi, Rumana Huque, Rusham Zahra Rana, Ruth Verhey, Saima Afaq, Sandro Rodrigues, Sesergio Baldew, Sushama Kanan, Yang William Zhao
`o` Agreement stages – Aishwarya Lakshmi Vidyasagaran, Carlos A Aguilar-Salinas, Corrado Barbui, Darwin Del Castillo, Eleonora Uphoff, Gerardo Zavala Gomez, Jaime Miranda, Jamie J Kirkham, Jan R Boehnke, Jessica Hanae Zafra Tanaka, John R Hurst, Jose Patricio López-Jaramillo, Kamran Siddiqi, Koralagamage Kavindu Appuhamy, Krishna Prasad Muliyala, Laura Downey, Mehreen Riaz Faisal, Najma Siddiqi, Naomi Levitt, Noemia Siqueira, Olga P García, Oscar Flores-Flores, Praveen Devarsetty, Rabeea Aman, Rajesh Vedanthan, Rakesh Singh, Richard IG Holt, Rubab Ayesha, Ruth Verhey, Sailesh Mohan, Saima Afaq, Syed Rahmat Ali, Yang William Zhao
`o` Data analysis – Aishwarya Lakshmi Vidyasagaran, Jan R Boehnke, Koralagamage Kavindu Appuhamy, Mehreen Riaz Faisal
`o` Data interpretation – Aishwarya Lakshmi Vidyasagaran, Jaime Miranda, Jamie Kirkham, Jan R Boehnke, John R Hurst, Kamran Siddiqi, Louise Rose, Mehreen Riaz Faisal, Najma Siddiqi, Rajesh Vedanthan
`o` Manuscript writing – Aishwarya Lakshmi Vidyasagaran, Jan R Boehnke, Louise Rose, Mehreen Riaz Faisal, Najma Siddiqi
`o` Revision of manuscript and editing – A Cristina Garcia-Ulloa, Aishwarya Lakshmi Vidyasagaran, Adewale L Oyeyemi, Asiful Haider Chowdhury, Augustine Nonso Odili, Carlos A Aguilar-Salinas, Cecilia Anza Ramirez, Corrado Barbui, Darwin Del Castillo, Dolores Ronquillo, Eleonora Uphoff, Gerardo Zavala Gomez, Gina Agarwal, Humaira Khalid, Jaime Miranda, Jamie J Kirkham, Jan R Boehnke, Jessica Hanae Zafra Tanaka, John R Hurst, Johnblack Kabukye, Jose Patricio López-Jaramillo, Josefien van Olmen, Joseph Senyo Kwashie, Judy Wright, Kamran Siddiqi, Kamrun N Koly, Koralagamage Kavindu Appuhamy, Kingsley Akinroye, Krishna Prasad Muliyala, Laura Downey, Louise Rose, María del Carmen Caamaño, Marianna Purgato, Mehreen Riaz Faisal, Najma Siddiqi, Naomi Levitt, Niels Victor Pacheco Barrios, Noemia Siqueira, Olga P García, Oscar Flores-Flores, Pervaiz Tufail, Phuong Tran Bich, Praveen Devarsetty, Qirat Naz, Rabeea Aman, Rajesh Vedanthan, Rakesh Singh, Richard IG Holt, Rubab Ayesha, Rufus Olusola Akinyemi, Rumana Huque, Rusham Zahra Rana, Ruth Verhey, Sailesh Mohan, Saima Afaq, Sandro Rodrigues, Sesergio Baldew, Sushama Kanan, Syed Rahmat Ali, Yang William Zhao

All authors had full access to all the data in the study and had final responsibility for the decision to submit for publication.

## COMPETING INTERESTS

None declared.

## FUNDING

This research was funded by the National Institute for Health Research (NIHR) Grants 17/63/130 NIHR Global Health Research Group: Improving Outcomes in Mental and Physical Multimorbidity and Developing Research Capacity (IMPACT) in South Asia and Grant NIHR203248 Global Health Centre for Improving Mental and Physical Health Together using UK aid from the UK Government to support global health research. The views expressed in this publication are those of the author(s) and not necessarily those of the NIHR or the UK government. Oscar Flores-Flores is supported by the Fogarty International Center and National Institute of Mental Health (NIMH) of the National Institutes of Health (NIH), United States under Award Number K43TW011586. The content is solely the responsibility of the authors and does not necessarily represent the official views of the National Institutes of Health.

This work was also supported by the Global Alliance for Chronic Diseases (GACD) Multimorbidity Subgroup and by members of the World Psychiatric Association Comorbidity Section. The GACD also part funded the DelphiManager software.

The NCD Alliance provided advice on engaging people with multimorbidity and publicised the study.

## Members of the ’COSMOS collaboration’ (collective authorship), in alphabetical order

1. Afaq, Saima; Khyber Medical University, Institute of Public Health and Social Sciences; Imperial College London, Department of Epidemiology and Biostatistics
2. Agarwal, Gina; McMaster University Faculty of Health Sciences, Family Medicine
3. Aguilar-Salinas, Carlos; Salvador Zubiran National Institute of Medical Sciences and Nutrition, Endocrinology and Metabolism
4. Akinroye, Kingsley; Nigerian Heart Foundation
5. Akinyemi, Rufus Olusola; University of Ibadan, Neuroscience and Ageing Research Unit, Institute for Advanced Medical Research and Training
6. Ali, Syed Rahmat; Khyber Medical University
7. Aman, Rabeea; Foundation University Islamabad
8. Appuhamy, Koralagamage Kavindu; University of York, Department of Health Sciences
9. Baldew, Se-Sergio; Anton de Kom University of Suriname, Physical Therapy Department
10. Barbui, Corrado; University of Verona, Department of Neuroscience, Biomedicine and Movement Sciences, Section of Psychiatry
11. Barrios, Niels Victor Pacheco; Universidad Peruana Cayetano Heredia, CRONICAS Center of Excellence in Chronic Diseases
12. Bich, Phuong Tran; University of Antwerp, Department of Family Medicine and Population Health
13. Caamaño, María del Carmen; Universidad Autónoma de Querétaro, School of Natural Sciences
14. Del Castillo Fernández, Darwin; Universidad Peruana Cayetano Heredia, CRONICAS Center of Excellence in Chronic Diseases
15. Haidar, Asiful; ARK foundation, Bangladesh,
16. Praveen, Devarsetty; The George Institute for Global Health India
17. Downey, Laura; UNSW; Imperial College London
18. Teixeira, Noemia; University of York, Department of Health Sciences
19. Flores-Flores, Oscar; Universidad de San Martin de Porres Facultad de Medicina Humana, Centro de Investigación del Envejecimiento (CIEN); Universidad Cientifica del Sur Facultad de Ciencias de la Salud
20. Gomez, Gerardo Zavala; University of York, Department of Health Sciences
21. Holt, Richard; University of Southampton, Human Development and Health, Faculty of Medicine
22. Huque, Rumana; ARK Foundation, Research and Development; University of Dhaka, Department of Economics
23. Kabukye, Johnblack; Uganda Cancer Institute; Stockholm University, SPIDER, Department of Computer and Systems Sciences
24. Kanan, Sushama; ARK foundation, Bangladesh
25. Khalid, Humaira; Pakistan Institute of Living and Learning
26. Koly, Kamrun; International Centre for Diarrheal Diseases Research, Bangladesh
27. Kwashie, Joseph Senyo; Community and Family Aid Foundation
28. Levitt, Naomi S.; University of Cape Town
29. Lopez-Jaramillo, Patricio; Universidad de Santander, Masira Research Institute, Medical School
30. Mohan, Sailesh; Public Health Foundation of India
31. García, Olga P.; Universidad Autónoma de Querétaro, School of Natural Sciences
32. Prasad-Muliyala, Krishna; National Institute of Mental Health and Neurosciences
33. Naz, Qirat; Benazir Bhutto Hospital, Institute of Psychiatry
34. Odili, Augustine; University of Abuja College of Health Sciences, Circulatory Health Research Laboratory
35. van Olmen, Josefien; University of Antwerp, Department of Primary and Interdisciplinary Care
36. Oyeyemi, Adewale; Arizona State University, College of Health Solutions; University of Maiduguri, Department of Physiotherapy
37. Purgato, Marianna; University of Verona, Neurosciences, Biomedicine and Movement Sciences
38. Zafra-Tanaka, Jessica; Universidad Peruana Cayetano Heredia, CRONICAS Center of Excellence in Chronic Diseases
39. Anza-Ramírez, Cecilia; Universidad Peruana Cayetano Heredia, CRONICAS Centre of Excellence in Chronic Diseases
40. Rodrigues Batista, Sandro Rogerio; Universidade Federal de Goiás, Family Medicine and Primary Health Care
41. Ronquillo, Dolores; Universidad Autónoma de Querétaro, School of Natural Sciences
42. Siddiqi, Kamran; University of York, Department of Health Sciences; Hull York Medical School
43. Singh, Rakesh; KIST Medical College, Department of Public Health
44. Tufail, Pervaiz; University of York, Department of Health Sciences
45. García Ulloa, Ana Cristina; Instituto Nacional de Ciencias Medicas y Nutricion Salvador Zubiran, CAIPaDi
46. Uphoff, Eleonora; University of York, Centre for Reviews and Dissemination
47. Verhey, Ruth; University of Zimbabwe, Research Support Centre, College of Health Sciences
48. Wright, Judy; University of Leeds Leeds Institute of Health Sciences
49. Zhao, Yang; Peking University Health Science Centre, The George Institute for Global Health; WHO, Collaborating Centre on Implementation Research for Prevention & Control of NCDs

## Non-author contributors

- Jibril Abdulmalik; Nigeria
- Anoshmita Adhikary; India
- Isaac Kwaku Adu; Ghana
- Helal Uddin Ahmed; Bangladesh
- Naveed Ahmed; Bangladesh
- Christel Antonius-Smits; Suriname
- Anas Ashraful; Bangladesh
- Faiza Aslam; UK
- Job Van Boven; New Zealand
- Camilla Cadorin; Italy
- Ram Krishna Chandyo; Nepal
- Alyssa Chase; UK
- Rachel Churchill; UK
- Meena Daivadanam; Sweden
- Anthony Danso-Appiah; Ghana
- Santa Kumar Das; Nepal
- Daniella Eiloof; Suriname
- Tonatiuh Barrientos Gutierrez; Mexico
- Najma Hayat; Pakistan
- Victoria Cavero Huapaya; UK
- Khaleda Islam; Bangladesh
- Abdul Kuddus; Bangladesh
- Saidur R Mashreky; Bangladesh
- Daniel Mograbi; Brazil
- Anum Naz; Pakistan
- Zara Nisar; UK
- Bruno P Nunes; Brazil
- Adesola Odole; Nigeria
- Obehi H Okojie; Nigeria
- Abdrahamane Ouedraogo; Burkina Faso
- Claudia Bambs Sandoval; Chile
- Arun Kumar Sharma; Nepal
- Sanjib Kumar Sharma; Nepal
- Elizabeth Shayo; Tanzania
- Abdou K Sillah; UK
- Lijing L. Yan; China
- Alejandro Zevallos-Morales; Peru

The authors extend their gratitude to all country partners, local researchers, and collaborators for their invaluable contributions in data collection, interpretation, transcription, and translation for this project.

## Notes

### Competing Interest Statement

The authors have declared no competing interest.

### Author Declarations

Health Sciences Research Governance Committee at the University of York (HSRGC/2020/409/D: COSMOS). Approvals were also obtained from relevant in-country ethics committees for all participating interview sites.

