## Appendices 1-6 for "Core outcome sets for trials of interventions to prevent and to treat multimorbidity in low- and middle-income countries: the COSMOS study"

### Appendix 1: The COS-STAR Statement Checklist

| **SECTION/TOPIC** | **ITEM** | **CHECKLIST ITEM** | **REPORTED ON PAGE NUMBER** |
| --- | --- | --- | --- |
| TITLE/ABSTRACT | | | |
| Title | 1a | Identify in the title that the paper reports the development of a COS | 1 |
| Abstract | 1b | Provide a structured summary | 1-2 |
| INTRODUCTION | | | |
| Background and Objectives | 2a | Describe the background and explain the rationale for developing the COS. | 4 |
|  | 2b | Describe the specific objectives with reference to developing a COS. | 4 |
| Scope | 3a | Describe the health condition(s) and population(s) covered by the COS. | 4 |
|  | 3b | Describe the intervention(s) covered by the COS. | 4 |
|  | 3c | Describe the setting(s) in which the COS is to be applied. | 4 |
| METHODS | | | |
| Protocol/Registry Entry | 4 | Indicate where the COS development protocol can be accessed, if available, and/or the study registration details. | 4 |
| Participants | 5 | Describe the rationale for stakeholder groups involved in the COS development process, eligibility criteria for participants from each group, and a description of how the individuals involved were identified. | 5-7, in different stages |
| Information Sources | 6a | Describe the information sources used to identify an initial list of outcomes. | 5, outcome generation stage |
|  | 6b | Describe how outcomes were dropped/combined, with reasons (if applicable). | 5, also figures 1a and 1b |
| Consensus Process | 7 | Describe how the consensus process was undertaken. | 6-7, agreement stage |
| Outcome Scoring | 8 | Describe how outcomes were scored and how scores were summarised. | 6-7, also Table 1 |
| Consensus Definition | 9a | Describe the consensus definition. | 7 |
|  | 9b | Describe the procedure for determining how outcomes were included or excluded from consideration during the consensus process. | 7 |
| Ethics and Consent | 10 | Provide a statement regarding the ethics and consent issues for the study. | 7 |
| RESULTS | | | |
| Protocol Deviations | 11 | Describe any changes from the protocol (if applicable), with reasons, and describe what impact these changes have on the results. | 11, discussion (not major) |
| Participants | 12 | Present data on the number and relevant characteristics of the people involved at all stages of COS development. | 8-10, in different stages |
| Outcomes | 13a | List all outcomes considered at the start of the consensus process. | Table 3 |
|  | 13b | Describe any new outcomes introduced and any outcomes dropped, with reasons, during the consensus process. | 9, also Table 3 |
| COS | 14 | List the outcomes in the final COS. | 1 (Abstract), 10 (Results), also Table 3 |
| DISCUSSION | | | |
| Limitations | 15 | Discuss any limitations in the COS development process. | 11-12 |
| Conclusions | 16 | Provide an interpretation of the final COS in the context of other evidence, and implications for future research. | 12 |
| OTHER INFORMATION | | | |
| Funding | 17 | Describe sources of funding/role of funders. | 21 |
| Conflicts of Interest | 18 | Describe any conflicts of interest within the study team and how these were managed. | 21 |

*From: Kirkham JJ, Gorst S, Altman DG, Blazeby JM, Clarke M, Devane D, et al. (2016) Core Outcome Set–STAndards for Reporting: The COS-STAR Statement. PLoS Med 13(10): e1002148. https://doi.org/10.1371/journal.pmed.1002148*

### Appendix 2: Appendices related to systematic review.

#### 2.1. Databases searched and example search strategy.

1. MEDLINE (Ovid) (see search strategy below)
2. Embase (Ovid)
3. PsycINFO (Ovid)
4. Cochrane Central Register of Controlled Trials (CENTRAL)
5. CINAHL (EBSCOhost)
6. Global Health (Ovid)
7. PakMediNet (<https://www.pakmedinet.com/>)
8. Global Index Medicus (WHO) (ISMEAR, LILACS, IMEMR, WRIM).
9. Cochrane Database of Systematic Reviews (CDSR)
10. The Database of Abstracts of Reviews of Effects (DARE)
11. PROSPERO (<https://www.crd.york.ac.uk/prospero/>)
12. Joanna Briggs
13. ClinicalTrials.gov
14. ISRCTN registry ([www.isrctn.com](http://www.isrctn.com))
15. The International Clinical Trials Registry Platform (<http://apps.who.int/trialsearch>).

Database: Ovid MEDLINE(R) ALL <1946 to July 17, 2020>

Search Strategy:

--------------------------------------------------------------------------------

1	Comorbidity/ (107657)

2	(comorbid$ or co-morbid$).ti,ab,kf. (181237)

3	(multimorbid$ or multi-morbid$).ti,ab,kf. (5739)

4	(multidisease$ or multi-disease$ or (multiple adj (ill$ or disease$ or condition$ or syndrome$ or disorder$))).ti,ab,kf. (5517)

5	((cooccur$ or co-occur$ or coexist$ or co-exist$ or multipl$) adj3 (disease$ or ill$ or care or condition$ or disorder$ or health$ or syndrome$)).ti,ab,kf. (76531)

6	1 or 2 or 3 or 4 or 5 (319021)

7	Cooperative Behavior/ (43408)

8	Interdisciplinary Communication/ (17155)

9	Interprofessional Relations/ (51072)

10	Intersectoral Collaboration/ (1958)

11	Patient Care Team/ (65264)

12	(collaborat$ adj4 care).ti,ab,kf. (8703)

13	((integrated or interdisciplinary or inter-disciplinary or interprofessional or inter-professional or intersectoral or inter-sectoral or multidisciplnary or multi-disciplinary) adj1 (care or management or practice or treatment)).ti,ab,kf. (12540)

14	7 or 8 or 9 or 10 or 11 or 12 or 13 (168849)

15	6 or 14 (482945)

16	(afghanistan or albania or algeria or american samoa or angola or "antigua and barbuda" or antigua or barbuda or argentina or armenia or armenian or aruba or azerbaijan or bahrain or bangladesh or barbados or republic of belarus or belarus or byelarus or belorussia or byelorussian or belize or british honduras or benin or dahomey or bhutan or bolivia or "bosnia and herzegovina" or bosnia or herzegovina or botswana or bechuanaland or brazil or brasil or bulgaria or burkina faso or burkina fasso or upper volta or burundi or urundi or cabo verde or cape verde or cambodia or kampuchea or khmer republic or cameroon or cameron or cameroun or central african republic or ubangi shari or chad or chile or china or colombia or comoros or comoro islands or iles comores or mayotte or democratic republic of the congo or democratic republic congo or congo or zaire or costa rica or "cote d’ivoire" or "cote d’ ivoire" or cote divoire or cote d ivoire or ivory coast or croatia or cuba or cyprus or czech republic or czechoslovakia or djibouti or french somaliland or dominica or dominican republic or ecuador or egypt or united arab republic or el salvador or equatorial guinea or spanish guinea or eritrea or estonia or eswatini or swaziland or ethiopia or fiji or gabon or gabonese republic or gambia or "georgia (republic)" or georgian or ghana or gold coast or gibraltar or greece or grenada or guam or guatemala or guinea or guinea bissau or guyana or british guiana or haiti or hispaniola or honduras or hungary or india or indonesia or timor or iran or iraq or isle of man or jamaica or jordan or kazakhstan or kazakh or kenya or "democratic people’s republic of korea" or republic of korea or north korea or south korea or korea or kosovo or kyrgyzstan or kirghizia or kirgizstan or kyrgyz republic or kirghiz or laos or lao pdr or "lao people's democratic republic" or latvia or lebanon or lebanese republic or lesotho or basutoland or liberia or libya or libyan arab jamahiriya or lithuania or macau or macao or "macedonia (republic)" or macedonia or madagascar or malagasy republic or malawi or nyasaland or malaysia or malay federation or malaya federation or maldives or indian ocean islands or indian ocean or mali or malta or micronesia or federated states of micronesia or kiribati or marshall islands or nauru or northern mariana islands or palau or tuvalu or mauritania or mauritius or mexico or moldova or moldovian or mongolia or montenegro or morocco or ifni or mozambique or portuguese east africa or myanmar or burma or namibia or nepal or netherlands antilles or nicaragua or niger or nigeria or oman or muscat or pakistan or panama or papua new guinea or new guinea or paraguay or peru or philippines or philipines or phillipines or phillippines or poland or "polish people's republic" or portugal or portuguese republic or puerto rico or romania or russia or russian federation or ussr or soviet union or union of soviet socialist republics or rwanda or ruanda or samoa or pacific islands or polynesia or samoan islands or navigator island or navigator islands or "sao tome and principe" or saudi arabia or senegal or serbia or seychelles or sierra leone or slovakia or slovak republic or slovenia or melanesia or solomon island or solomon islands or norfolk island or norfolk islands or somalia or south africa or south sudan or sri lanka or ceylon or "saint kitts and nevis" or "st. kitts and nevis" or saint lucia or "st. lucia" or "saint vincent and the grenadines" or saint vincent or "st. vincent" or grenadines or sudan or suriname or surinam or dutch guiana or netherlands guiana or syria or syrian arab republic or tajikistan or tadjikistan or tadzhikistan or tadzhik or tanzania or tanganyika or thailand or siam or timor leste or east timor or togo or togolese republic or tonga or "trinidad and tobago" or trinidad or tobago or tunisia or turkey or "turkey (republic)" or turkmenistan or turkmen or uganda or ukraine or uruguay or uzbekistan or uzbek or vanuatu or new hebrides or venezuela or vietnam or viet nam or middle east or west bank or gaza or palestine or yemen or yugoslavia or zambia or zimbabwe or northern rhodesia or global south or africa south of the sahara or sub-saharan africa or subsaharan africa or africa, central or central africa or africa, northern or north africa or northern africa or magreb or maghrib or sahara or africa, southern or southern africa or africa, eastern or east africa or eastern africa or africa, western or west africa or western africa or west indies or indian ocean islands or caribbean or central america or latin america or "south and central america" or south america or asia, central or central asia or asia, northern or north asia or northern asia or asia, southeastern or southeastern asia or south eastern asia or southeast asia or south east asia or asia, western or western asia or europe, eastern or east europe or eastern europe or developing country or developing countries or developing nation? or developing population? or developing world or less developed countr* or less developed nation? or less developed population? or less developed world or lesser developed countr* or lesser developed nation? or lesser developed population? or lesser developed world or under developed countr* or under developed nation? or under developed population? or under developed world or underdeveloped countr* or underdeveloped nation? or underdeveloped population? or underdeveloped world or middle income countr* or middle income nation? or middle income population? or low income countr* or low income nation? or low income population? or lower income countr* or lower income nation? or lower income population? or underserved countr* or underserved nation? or underserved population? or underserved world or under served countr* or under served nation? or under served population? or under served world or deprived countr* or deprived nation? or deprived population? or deprived world or poor countr* or poor nation? or poor population? or poor world or poorer countr* or poorer nation? or poorer population? or poorer world or developing economy* or less developed economy* or lesser developed economy* or under developed economy* or underdeveloped economy* or middle income economy* or low income economy* or lower income economy* or low gdp or low gnp or low gross domestic or low gross national or lower gdp or lower gnp or lower gross domestic or lower gross national or lmic or lmics or third world or lami countr* or transitional countr* or emerging economies or emerging nation?).ti,ab,sh,kf. (1919265)

17	(afghan or afghans or afghani or albanian? algerian? or american samoan? or angolan? or antiguan? or barbudan? or argentine? or argentinian? or argentinean? or armenian? or aruban? or azerbaijani? or bahraini? or bangladeshi? or bangalees or bajan? or belarusian? or byelorussian? or belizean? or beninese? or bhutanese or bolivian? or bosnian? or botswana or batswana or brazilian? or brasilian? or bulgarian? or burkinabe or burkinese or burundian? or cape verdean? or cabo verdean? or cambodian? or khmer or cameroonian? or central african? or chadian? or chilean? or chinese or colombian? or comorian? or congolese or costa rican? or ivorian? or croatian? or cuban? or cypriot? or czech? or djiboutian? or dominican? or ecuadorian? or egyptian? or salvadoran? or equatorial guinean? or equatoguinean? or eritrean? or estonian? or swazi? or swati? or ethiopian? or fijian or gabonese or gabonaise or gambian? or georgian? or ghanaian? or gibraltarian? or greek? or grenadian? or guamanian? or guatemalan? or guinean? or bissau guinean? or guyanese or haitian? or honduran? or hungarian? or indian? or indonesian? or iranian? or iraqian? or iraqi? or manx or jamaican? or jordanian? or kazakhstani? or kenyan? or kirabati or kirabatian? or north korean? or korean? or kosovar? or kosovan? or kyrgyz* or lao or laotian? or latvian? or lebanese or lesothan? or lesothonian? or mosotho or basotho or liberian? or libyan? or lithuanian? or macanese or macedonian? or malagasy or madagascan? or malawian? or malaysian? or maldivian? or malian? or maltese or marshallese? or mauritanian? or mauritian? or mexican? or micronesian? or moldovan? or mongolian? or mongol or montenegrin? or moroccan? or mozambican? or burmese or myanma or namibian? or nauruan? or nepali or nepalese or netherlands antillean? or nicaraguan? or nigerien? or nigerian? or northern mariana islander? or mariana? or omani? or pakistani? or palauan? or panamanian? or papua new guinean? or paraguayan? or peruvian? or philippine? or philipine? or phillipine? or phillippine? or filipino? or filipina? or polish or pole or poles or portuguese or puerto rican? or romanian? or russian? or soviet people or soviet population or rwandan? or rwandese or ruandan? or ruandese or samoan? or sao tomean? or santomean? or saudi arabian? or saudi? or senegalese or serbian? or montenegrin? or seychellois or seychelloise? or sierra leonean? or slovak? or slovene? or solomon islander? or somali? or south african? or south sudanese or sri lankan? or ceylonese or kittitian? or nevisian? or saint lucian? or vincentian? or sudanese or surinamese? or syrian? or tajik? or tajikistani? or tanzanian? or tanganyikan? or thai or timorese? or togolese or tongan? or trinidadian? or tobagonian? or tunisian? or turk? or turkish or turkmen? or tuvaluan? or ugandan? or ukrainian? or uruguayan? or uzbek? or vanuatu* or venezuelan? or vietnamese or yemeni? or yemenite? or yemenese or yugoslav? or yugoslavian? or zambian? or zimbabwean?).ti,ab,sh,kf. (840691)

18	16 or 17 (2283252)

19	randomized controlled trial.pt. (509754)

20	controlled clinical trial.pt. (93762)

21	randomized.ab. (486234)

22	randomised.ab. (97082)

23	placebo.ab. (209447)

24	clinical trials as topic.sh. (192103)

25	randomly.ab. (337079)

26	trial.ti. (221842)

27	19 or 20 or 21 or 22 or 23 or 24 or 25 or 26 (1337740)

28	exp animals/ not humans.sh. (4718707)

29	27 not 28 (1233028)

30	15 and 18 and 29 (2569)

31	exp Child/ (1906042)

32	exp Adult/ (7189354)

33	31 not (31 and 32) (1210187)

34	30 not 33 (2343)

35	limit 34 to yr="1990 -Current" (2335)

***************************

#### 2.2. PRISMA flow diagram

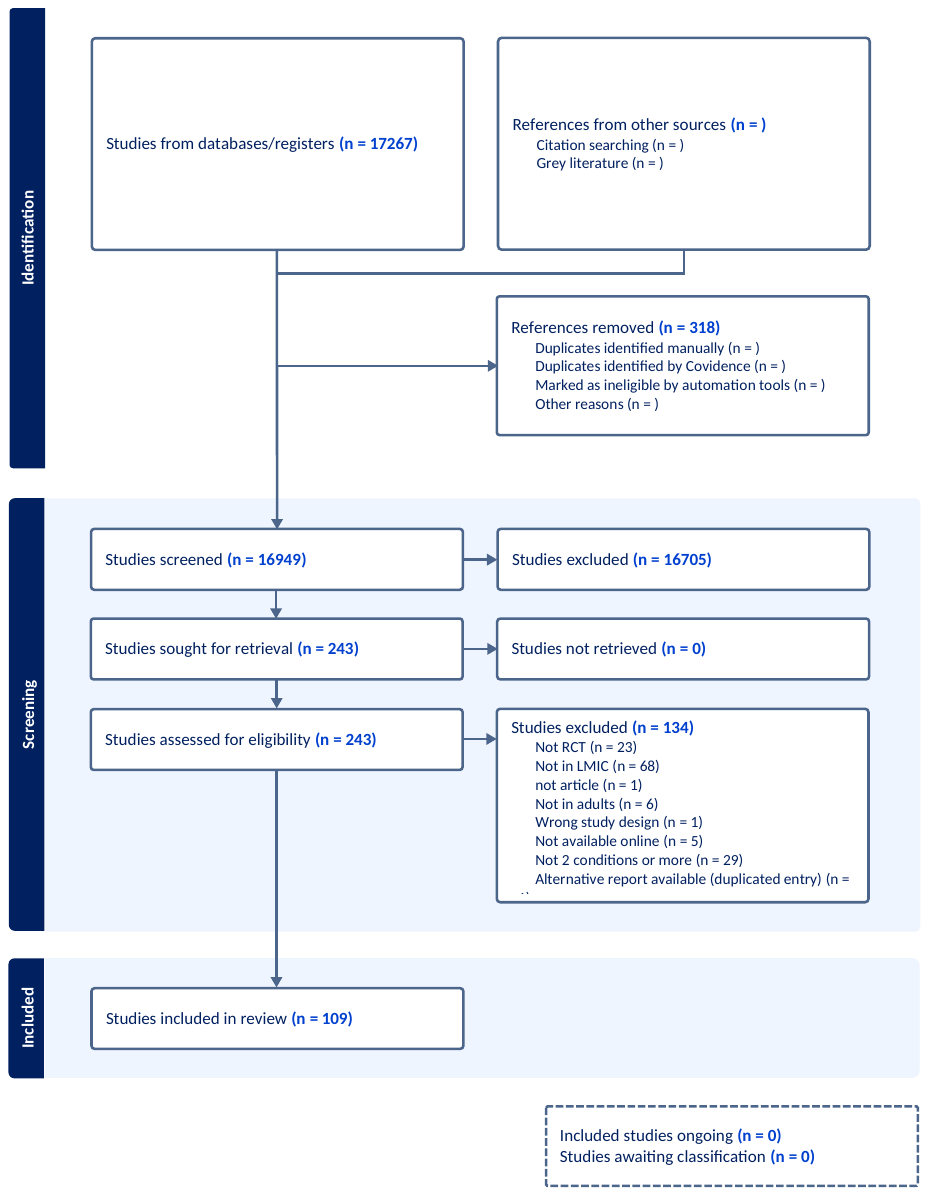

#### 2.3. Table of characteristics of included studies

| **Study ID** | **Title** | **Country** | **Target conditions** |
| --- | --- | --- | --- |
| Wang 2014 | Effect of amlodipine plus atorvastatin calcium on hypertension patients complicated with hyperlipidaemia | China | Hypertension and hyperlipidaemia |
| Sanjeev 2013 | Nevirapine vs. efavirenz-based ART regimens in antiretroviral-naive patients with HIV and tuberculosis infections in India: a pilot study | India | HIV and TB |
| Liang 2013 | Effect of laparoscopic Roux-en-Y gastric bypass surgery on type 2 DM with hypertension: a randomized controlled trial | China | T2DM, obesity, hypertension |
| Li 2015 | Observation of efficacy of trimetazidine in treatment of CHD combined with diabetes | China | Coronary heart disease and diabetes |
| Wang 2020 | A Chinese herbal formula shows beneficial effects on comorbid depression and coronary heart disease based on the philosophy of psycho-cardiology | China | Depression and Coronary Heart disease |
| Xinju 2017 | The efficacy of internet-based cognitive behaviour therapy on blood pressure for comorbid hypertension and insomnia | China | Hypertension and insomnia |
| Xin 2002 | A Clinical Trial of Paroxetine and Psychotherapy in Patients with Poststroke Depression and Anxiety | China | Acute brain stroke, depression, anxiety |
| Freitas Júnior 2010 | Estudo comparativo dos efeitos agudos do sildenafil e nitroprussiato de sódio sobre a hipertensão pulmonar de pacientes com insuficiência cardíaca avançada | Brazil | Pulmonary hypertension and heart failure |
| Duarte 2011 | Síndrome da apnéia do sono na acromegalia: impacto do tratamento sobre o metabolismo dos carboidratos | Brazil | Acromegaly and sleep apnoea |
| Mokhber 2011 | [Comparison of desipramine and sertraline in the treatment of depression in patients suffering from Alzheimer disease] | Iran | Depression in Alzheimer's patients |
| Morad Rasouli 2009 | [Effects of cognitive behavioural group therapy in male opioid dependent patients comorbid with major depressive disorder] | Iran | Depression in opioid-dependent men receiving methadone treatment |
| Huiying 2012 | Influence of comprehensive nursing intervention on pulmonary function and depressive mood of patients with COPD and depression | China | COPD and depression |
| Bündchen 2010 | Ausência de influência da massa corporal na redução da pressão arterial após exercício físico | Brazil | Hypertension and Overweight or Obesity |
| An-Lin 2006 | Effects of antidepressant therapy in patients with suspected angina pectoris and negative coronary angiogram complicating comorbid depression | China | Suspected angina pectoris with negative coronary angiogram and depression |
| Aiyun 2017 | A study on the treatment of depression and anxiety in elderly patients by Shu gan jie yu capsule combined with Ascitalopram | China | Anxiety, Depression |
| Buranakitjaroen 2007 | Efficacy and safety of sildenafil in Asian males with erectile dysfunction and cardiovascular risk | Thailand, Malaysia | Erectile dysfunction, with one or more of HT, dyslipidaemia, T2DM |
| Vanelle 2006 | A double-blind RCT of amisulpride versus olanzapine for 2 months in the treatment of subjects with schizophrenia and comorbid depression | France, Italy, and Tunisia. | Schizophrenia and Comorbid depression |
| Dong 2005 | Anxiolytic intervention for coronary heart disease patients combined with anxiety disorder. [Chinese] | China | coronary heart disease and anxiety |
| Manosuthi 2005 | Efavirenz levels and 24-week efficacy in HIV-infected patients with tuberculosis receiving highly active ART and rifampicin | Thailand | HIV and TB |
| Liu 2005 | Effect and safety of the treatment for comorbid anxiety and depression by use of short-term and low-dose olanzapine in combination with fluoxetine. | China | Anxiety and depression |
| Emre 2004 | Rivastigmine for dementia associated with Parkinson's disease | 12 countries | Dementia and Parkinsons |
| Miti 2003 | Integration of TB treatment in a community-based home care programme for persons living with HIV/AIDS in Ndola, Zambia | Zambia | HIV and TB |
| Li 2009 | Beneficial impact of Xuezhikang on cardiovascular events and mortality in elderly hypertensive patients with previous myocardial infarction from the China Coronary Secondary Prevention Study (CCSPS) | China (19 provinces) | HT and Myocardial inaction |
| NCT 2013 | Chinese Older Adults-Collaboration in Health (COACH)Study | China | Hypertension and depression |
| Poirier 2009 | Adding prompt revascularization to medical therapy did not reduce mortality or CV events in patients with type 2 diabetes and CAD | 6 countries | Type 2 diabetes and heart disease |
| Moosa 2012 | Antidepressants versus interpersonal psychotherapy in treating depression in HIV-positive patients | South Africa | Depression in HIV patients receiving antiretroviral treatment |
| PrayGod 2012 | The effect of energy-protein supplementation on weight, body composition and handgrip strength among pulmonary tuberculosis HIV-co-infected patients: randomised controlled trial in Mwanza, Tanzania | Tanzania | TB and HIV |
| Greenfield 2010 | Integrated Management of Physician-delivered Alcohol Care for Tuberculosis Patients: Design and Implementation | Russia | Alcohol Use in people with TB |
| Nyamathi 2012 | Impact of an ASHA intervention on depressive symptoms among rural women living with AIDS in India | India | Depression in women living with AIDS |
| Durovni 2010 | The implementation of isoniazid preventive therapy in HIV clinics: the experience from the TB/HIV in Rio (THRio) study | Brazil | TB in HIV patients |
| Shah 2012 | Improving diabetes care: Multi-component cardiovascular disease risk reduction strategies for people with diabetes in South Asia: CARRS | India and Pakistan | Risk of CVD in patients with Diabetes |
| Heeley 2011 | The Sleep Apnoea cardioVascular Endpoints (SAVE) study: 1000 recruited patients | 5 countries | OSA, cardiovascular disease |
| Peltzer 2013 | Screening and brief interventions for hazardous and harmful alcohol use among patients with active tuberculosis attending primary public care clinics in South Africa: results from a cluster randomized controlled trial | South Africa | Alcohol misuse among active TB patients |
| Palavras 2015 | Comparing cognitive behavioural therapy for eating disorders integrated with behavioural weight loss therapy to cognitive behavioural therapy-enhanced alone in overweight or obese people with bulimia nervosa or binge eating | Brazil | Bulimia nervosa/eating disorders and overweight/obesity |
| NCT 2016a | Diabetes Complication Control in Community Clinics (D4C) Trial | China | CVD risk in patients with diabetes |
| Fairall 2016 | Educational Outreach with an Integrated Clinical Tool for Nurse-Led Non-communicable Chronic Disease Management in Primary Care in South Africa: A Pragmatic Cluster Randomised Controlled Trial | South Africa | Hypertension, Diabetes, Chronic respiratory disease, Depression |
| NCT 2015 | The Effect of N- Acetylcysteine on Inflammatory and Oxidative Stress Biomarkers | Brazil | Bipolar Disorder |
| NCT 2014a | Effect of Supplementary Vitamin D in Patients With Diabetes Mellitus and Pulmonary Tuberculosis | Pakistan | Diabetes and Pulmonary TB |
| NCT 2016b | Effectiveness of a Mobile Technology Intervention for the Treatment of Depression | Brazil | Depressive symptoms and co-morbid diabetes and/or hypertension |
| Asl 2014 | Effectiveness of Mindfulness-Based Cognitive Therapy for Co-Morbid Depression in Drug-Dependent Males | Iran | Depression in drug dependent population |
| NCT 2016c | Effectiveness of Nurse-delivered Care for Adherence/Mood in HIV in South Africa | South Africa | HIV and Depressive Symptoms |
| Da Silva 2015 | Effects of exercise training and photobiomodulation therapy (EXTRAPHOTO) on pain in women with fibromyalgia and temporomandibular disorder: protocol for RCT | Brazil | Women with fibromyalgia and temporomandibular disorder |
| Pervane Vural 2016 | Effects of Mirror Therapy in Stroke Patients With Complex Regional Pain Syndrome Type 1: A Randomized Controlled Study | Turkey | First-time stroke and simultaneous complex regional pain syndrome |
| NCT 2014b | Improving Mental Health Through Integration With Primary Care in Rural Karnataka | India | Depression or anxiety and co-morbid diabetes, or cardiovascular disease |
| Hasanpour-Dehkordi 2016 | Effects of yoga on physiological indices, anxiety and social functioning in multiple sclerosis patients: a randomized trial | Iran | Multiple sclerosis (MS), Anxiety |
| Huo 2015 | Efficacy of folic acid therapy in primary prevention of stroke among adults with hypertension in China: The CSPPT randomized clinical trial | China | Stroke (prevention of stroke), Hypertension |
| Safi 2015 | A Fresh Look at the Mechanisms of Progressive Muscle Relaxation on Depression in Female Patients with Multiple Sclerosis | Iran | Depression, Multiple sclerosis (MS) |
| Petersen 2014 | A group-based counselling intervention for depression comorbid with HIV/AIDS using a task shifting approach in South Africa | South Africa | Depression and HIV/AIDS |
| Basheti 2016 | Home medication management review in outpatients with chronic diseases in Jordan: a randomized control trial | Jordan | Mean (SD) number of conditions per patient was 3.97 (1.56) |
| Song 2015 | Influence of different exercise nursing intervention plans on blood glucose level of type 2 DM patients complicated with obesity | China | Type 2 diabetes mellitus (T2DM) and obesity |
| NCT 2016d | Integrated Primary Care for Diabetes and Cardiovascular Disease | Brazil | Cardiovascular disease and diabetes |
| Wagner 2014 | INtegration of DEPression Treatment into HIV Care in Uganda (INDEPTH-Uganda): study protocol for a randomized controlled trial | Uganda | HIV and depression |
| Hu 2015 | Suan zao ren tang in combination with zhi zi chi tang as a treatment protocol for insomniacs with anxiety: parallel-controlled RCT | China | Insomnia and anxiety |
| Bolton 2014 | A transdiagnostic community-based mental health treatment for comorbid disorders: development and outcomes of a randomized controlled trial among Burmese refugees in Thailand | Thailand | Mental Health and Alcohol Use |
| Budnevsky 2016 | [Pulmonary rehabilitation as an effective method for optimizing therapeutic and preventive measures in patients with chronic obstructive pulmonary disease concurrent with metabolic syndrome] | Russia | Chronic obstructive pulmonary disease (COPD) concurrent with metabolic syndrome (MS) |
| Liu 2015 | Pulsed electromagnetic fields for postmenopausal osteoporosis and concomitant lumbar osteoarthritis in southwest China using proximal femur bone mineral density as the primary endpoint: Study protocol for a randomized controlled trial | China | Postmenopausal osteoporosis, lumbar osteoarthritis |
| Gurevich 2015 | [Psychotherapy in patients with alcohol dependence and comorbid endogenous pathology] | Russia | Alcohol dependence, developed against paranoid schizophrenia |
| Sevostyanova 2018 | [Combined rehabilitation in the patients presenting with dorsopathies of the lumbar spine and concomitant ibs based at a therapeutic clinic] | Russia | Dorsopathy of the lumbar spine, combined with IBS |
| Majeed 2018 | Bacillus coagulans MTCC 5856 for the management of major depression with irritable bowel syndrome: A randomised, double-blind, placebo controlled, multi-centre, pilot clinical study | India | Irritable bowel syndrome (IBS), Major depressive disorder (MDD) |
| Thapinta 2017 | Cognitive Behaviour Therapy Self-Help Booklet to Decrease Depression and Alcohol Use among People with Alcohol Dependence in Thailand | Thailand | Depression, Alcohol dependence |
| NCT 2018a | Cognitive Processing Therapy Versus Medication for the Treatment of Post Traumatic Stress Disorder & Substance Use Disorder Egyptian Patients | Egypt | Substance Use Disorder, Post-Traumatic Stress Disorder |
| Fairall 2018 | Collaborative care for the detection and management of depression among adults receiving antiretroviral therapy in South Africa: study protocol for the CobALT randomised controlled trial | South Africa | depression among HIV patients receiving antiretroviral therapy |
| Petersen 2018 | Collaborative care for the detection and management of depression among adults with hypertension in South Africa: protocol for the PRIME-SA RCT | South Africa | Depression and hypertension |
| Myers 2018 | Comparing dedicated and designated models of integrating mental health into chronic disease care: study protocol for a cluster RCT | South Africa | Alcohol misuse or depression in diabetic or HIV patients |
| NCT 2017a | Effect of Different Exercises on Musculoskeletal Pain, Glucose Level and Quality of Life Among Patients With Diabetes | South Africa | Diabetes, Depression |
| NCT 2018b | Effect of Dulaglutide on Liver Fat in Patients With Type 2 Diabetes and Nonalcoholic Fatty Liver Disease | India | Non-Alcoholic Fatty Liver Disease, Type 2 Diabetes Mellitus |
| Zhang 2017 | Effect of HBM Rehabilitation Exercises on Depression, Anxiety and Health Belief in Elderly Patients with Osteoporotic Fracture | China | Depression, anxiety in elderly with osteoporotic fracture |
| Tu 2018 | Hypertension management for community-dwelling older people with diabetes in Nanchang, China: study protocol for a cluster randomized controlled trial | China | Hypertension in older people with type 2 diabetes |
| Abas 2018 | Feasibility and acceptability of a task-shifted intervention to enhance adherence to HIV medication and improve depression in people living with HIV in Zimbabwe, a low-income country in sub-Saharan Africa | Zimbabwe | HIV and depression |
| Valiani 2018 | The effect of auriculotherapy on stress, anxiety, and depression in MS patients: A double blind randomized clinical control trial (parallel design) | Iran | Multiple sclerosis (MS), Anxiety, Depression |
| NCT 2017b | Effectiveness of a mHealth Intervention for the Treatment of Depression in People with Diabetes or Hypertension in Peru | Peru | Depression, Diabetes, Hypertension |
| Ahmadi 2018 | The effectiveness of different singly administered high doses of buprenorphine in reducing suicidal ideation in acutely depressed people with co-morbid opiate dependence: double-blind RCT | Iran | People living with severe Opioid Use Disorder and depression |
| Bahrani 2017 | The effectiveness of mindfulness-integrated cbt on depression, anxiety, and stress in females with multiple sclerosis: A single blind RCT | Iran | Multiple Sclerosis with Depression, Anxiety, and 'Stress' |
| Rashidi 2018 | The Effects of Acute Multisession Bifrontal Transcranial Direct Current Stimulation on Intractable Tinnitus and comorbid depression and anxiety: A Study Protocol for Randomized Controlled Trial | Iran | Intractable Tinnitus and comorbid depression and anxiety |
| Djazayery 2018 | The effects of alcoholic extract of saffron (Crocus satious L.) on mild to moderate comorbid depression-anxiety, sleep quality, and life satisfaction in type 2 diabetes mellitus: A double-blind, placebo-controlled RCT | Iran | Type 2 diabetic patients suffering from mild to moderate anxiety and depression |
| Moazen-Zadeh 2018 | Effects of Saffron on Cognition, Anxiety, and Depression in Patients Undergoing Coronary Artery Bypass Grafting: A Double-Blind Placebo-RCT | Iran | Depression and anxiety in patients undergoing CABG |
| Sudfeld 2017 | Efficacy of vitamin D3 supplementation in reducing incidence of pulmonary tuberculosis and mortality among HIV-infected Tanzanian adults initiating antiretroviral therapy: study protocol for a randomized controlled trial | Tanzania | pulmonary tuberculosis among HIV-infected |
| Noroozi 2017 | Improving Depression, and Quality of Life in Patients with Type 2 Diabetes: Using Group Cognitive Behavior Therapy | Iran | Type 2 diabetes, Depression |
| Bongomin 2018 | Integrated versus referred management of cardiovascular disease (CVD) risk factors for HIV-positive patients on antiretroviral therapy in Swaziland | Swaziland (Eswatini) | HIV, Cardiovascular disease risk factors |
| Keshavarz 2018 | Omega-3 supplementation effects on body weight and depression among dieter women with co-morbidity of depression and obesity compared with the placebo: A randomized clinical trial | Iran | depression and obesity |
| Kowalski 2017 | The INtegrating DEPrEssioN and Diabetes treatmENT (INDEPENDENT) study: Design and methods to address mental healthcare gaps in India | India | Depression and Diabetes |
| ISRCTN 2017 | An Intervention Program to Reduce the Risk Factors of Metabolic Syndrome in Malaysian Female University Staff with Polycystic Ovarian Syndrome | Malaysia | Polycystic Ovarian Syndrome (PCOS), Metabolic Syndrome |
| Steinert 2017 | Resource activation for treating post-traumatic stress disorder, co-morbid symptoms and impaired functioning: a RCT in Cambodia | Cambodia | Post-traumatic stress disorder (PTSD), Anxiety, Depression |
| Zemestani 2017 | A Preliminary Investigation on the Effectiveness of Unified and Transdiagnostic CBT for Patients with Comorbid Depression and Anxiety | Iran | Depression and anxiety |
| Jha 2017 and  Prabhakaran 2018 | Protocol for the mWellcare trial: a multicentre, cluster RCT to compare the effectiveness of mWellcare, an mHealth system for an integrated management of patients with hypertension and diabetes, versus EUC | India | Hypertension and diabetes |
| Chen 2018 | Protocol of an ongoing randomized controlled trial of care management for comorbid depression and hypertension: the Chinese Older Adult Collaborations in Health (COACH) study | China | depression and hypertension |
| Miller 2018 | A scalable, integrated intervention to engage people who inject drugs in HIV care and medication-assisted treatment (HPTN 074): a RCT phase 3 feasibility and efficacy study | Ukraine; Vietnam; Indonesia | People who inject drugs: HIV-infected and their HIV-uninfected injection partners. |
| ReBEC 2018 | Vitamin D action in the treatment of Depression and reduction of Cardiovascular Risk | Brazil | Depression, Cardiovascular risk |
| Caetano 2018 | Trial-Based Cognitive Therapy: Efficacy of a New CBT Approach for Treating Social Anxiety Disorder with Comorbid Depression | Brazil | Generalized social anxiety disorder in comorbid depression |
| ISRCTN 2019a | A collaborative care psychosocial intervention for lowering blood pressure among old adults with hypertension and depression in primary care in Guarulhos, Brazil: the PROACTIVE-H study | Brazil | Hypertension and depression in older adults. |
| Kane 2020 | Common Elements Treatment Approach (CETA) for unhealthy alcohol use among persons with HIV in Zambia: Study protocol | Zambia | HIV and high-risk alcohol use and possible mental health |
| Furtado 2019 | Dapagliflozin and Cardiovascular Outcomes in Patients with Type 2 Diabetes Mellitus and Previous Myocardial Infarction | 33 countries | Type II DM and those with previous MI |
| Khan 2019 | Efficacy of eye movement desensitization & reprocessing versus cognitive behavioural therapy in PTSD Symptoms: Study protocol for RCT | Pakistan | PTSD and depressive symptoms |
| Pirnia 2019 | Evaluation of the effectiveness of auricular acupuncture in suicidal ideation and cortisol level in dysthymic patients with comorbid opiate use | Iran | Dysthymic patients with comorbid opiate use disorders |
| ISRCTN 2019b | Determination of the Effect of Crude Oil on Erectile Dysfunction in Men with Multiple Sclerosis | Iran | Men with multiple sclerosis and erectile dysfunction |
| Li 2019 | Effect of a patient education and rehabilitation program on anxiety, depression and quality of life in muscle invasive bladder cancer patients | China | Anxiety, depression, and bladder cancer |
| Abdollahi 2020 | The effect of aromatherapy with bitter orange (Citrus aurantium) extract on anxiety and fatigue in type 2 diabetic patients | Iran | anxiety and fatigue in people with type 2 diabetes |
| Levy 2019 | Implementation research for public sector mental health care scale-up (SMART-DAPPER): a sequential multiple, assignment randomized trial | Kenya | Major depressive disorder and PTSD |
| Negaresh 2019 | Effect of Short-Term Interval Exercise Training on Fatigue, Depression, and Fitness in Normal Weight vs. Overweight Person with Multiple Sclerosis | Iran | Overweight and Multiple sclerosis |
| Zemestani 2020 | Effectiveness of mindfulness-based cognitive therapy for comorbid depression and anxiety in pregnancy: a randomized controlled trial | Iran | Comorbid depression and anxiety (in pregnant women) |
| Cruz 2019 | Morbidity in patients with uncontrolled hypertension and obstructive sleep apnoea: baseline profile of the MORPHEOS study | Brazil | Uncontrolled hypertension and Significant sleep apnoea |
| Mutyambizi-Mafunda 2019 | Integrating a brief mental health intervention into primary care services for patients with HIV and diabetes in South Africa: study protocol | South Africa | Diabetes, HIV, Depression, Hazardous alcohol use |
| Becerril-Alarcon 2019 | Inulin Supplementation Reduces Systolic Blood Pressure in Women with Breast Cancer Undergoing Neoadjuvant Chemotherapy | Mexico | Blood pressure in women with breast cancer receiving chemotherapy |
| Joska 2020 | Nurse-Delivered Cognitive Behavioral Therapy for Adherence and Depression Among People Living With HIV (the Ziphamandla Study): Protocol for a Randomized Controlled Trial | South Africa | HIV and depression |
| Citrome 2019 | Olanzapine Plus Samidorphan in Subjects with Schizophrenia and Comorbid Alcohol Use Disorder: Rationale and Design for a Phase II rct | US, Bulgaria, Poland | Schizophrenia, Alcohol Use Disorder |
| Akhondzadeh 2020 | A placebo controlled RCT of Crocus sativus L. (saffron) on depression and food craving among overweight women with mild to moderate depression | Iran | depression and obesity (in women) |
| Tu 2020 | A transitional care intervention for hypertension control for older people with diabetes: A cluster randomized controlled trial | China | Hypertension control in older people with diabetes |
| NCT 2019 | Vortioxetine vs Sertraline in Mexicans | Mexico | Type II diabetes and Depression |

### Appendix 3: Ethics committee approvals

| **Organisation** | **Country** | **Approving committee/ department** |
| --- | --- | --- |
| University of York, York | United Kingdom | Health Sciences Research Governance Committee (HSRGC/2020/409/D: COSMOS) |
| Afghan Thoughts for Development and Change, Kabul | Afghanistan | Department of Research and Development |
| Bangladesh University of Health Sciences, Dhaka | Bangladesh | Department of Public Health Sciences, Centre for Injury Prevention and Research Bangladesh |
| Association Mahidol de Sante Publique, Ouagadougou | Burkina Faso | Comite D'éthique Pour La Recherche En Sante |
| Community and Family Aid Foundation Ghana, Newtown-Accra | Ghana | CAFAF/20/COSMOS /3 (Local Reference: Project 101/20) |
| Universidad Autónoma de Querétaro, Santiago de Querétaro | Mexico | C.U., 1 de febrero de 2021 DIP/56-2021 |
| National Medical College, Birgunj | Nepal | Institutional Review Committee (IRC), Ref. F-NMC/518/077-078 |
| University of Benin Teaching Hospital, Benin City | Nigeria | Health Research Ethics Committee, ADM/E 22/A/VOL. VII/14830970 |
| Rawalpindi Medical University & Allied Hospitals, Rawalpindi | Pakistan | Research and Ethical Committee, 247/IREF/RMU/2020 |
| PRISMA ONG, Lima | Peru | Comité Institucional de Ética en Investigación, CE0981.20 |
| Ministerie van Volksgezondheid in Suriname | Suriname | Direction Centrale Administratie |

### Appendix 4: Examples codes for outcomes added from qualitative interviews.

| **Country** | **Quoted passage** | **Extracted outcome** |
| --- | --- | --- |
| Afghanistan | In my country, there are also limited health care providers who can rightly diagnose and treat these conditions. These all challenges made me worried about my health conditions. | Healthcare access |
| Afghanistan | Follow the doctor’s advice and keep the patient tracking his medication and treatment plan. | Adherence to treatment |
| Bangladesh | a lot of things were restricted, like food. | Diet |
| Bangladesh | encourage him emotionally to live a happy and healthy life. | Psychological wellbeing |
| Burkina Faso | So, I am more afraid. | Health anxiety |
| Burkina Faso | The impact for us is the high cost of care. | Healthcare cost |
| Ghana | We want treatment to help him heal and resume his normal activities. | Functioning/ADL |
| Ghana | I could not sleep. | Sleep |
| Mexico | And now I have my three haemodialysis sessions, and the truth, the haemodialysis, the diet, and exercise, have lifted me up in a phenomenal way. | Perceived health |
| Mexico | I hope that, well, I have a better life, stability | Health-related quality of life |
| Nepal | Accessibility of check-up in every household for screening such disease. | Timely screening |
| Nepal | She does not want to spend on me. My son sends money from abroad to daughter-in-law, but she doesn’t want to spend those on my treatment. | Carer burden |
| Nigeria | I’ve not been taking it much, that I take you see at time I don’t take it every time. | Adherence to treatment |
| Nigeria | So he now changed my drugs and then I was now complaining that the thing became severe. | Perceived health |
| Pakistan | Now she gets angry. | Agitation |
| Pakistan | Good quality medicines should be provided and used before, during and after treatment for both rich and poor without any discrimination. | Healthcare quality |
| Peru | I would hope the best not, to be able to live normal, to be able to make normal life. | Health-related quality of life |
| Peru | the CT scan was one hundred [soles] and so many, and the other was almost 300 soles, so at that time I did not have… | Healthcare cost |
| Suriname | She does not want additional medication prescribed or that the doses be increased. | Reduced medication |
| Suriname | Physical therapist taught me was to accept my disabilities. It was hard because I realized that my life would never be the same. | Acceptance of illness |

### Appendix 5: Prevention and Treatment outcomes from outcome generation and Delphi R1 stages, classified using Dodd’s taxonomy.

| **S no.** | **Outcome** | **Help text** | **Core areas*** | **Williamson/Clarke (revised scale) *** |
| --- | --- | --- | --- | --- |
| **Prevention outcomes** | | | | |
| 1 | Adherence to treatment | The extent to which a person keeps to a recommended treatment, such as taking prescribed drugs or following a diet or exercise programme | Life impact | Delivery of Care |
| 2 | Adverse events | Any unexpected or unintended negative consequences of an intervention, for example, allergic reaction to a drug, or injury from a recommended exercise programme | Adverse events | Adverse Events/ Effects |
| 3 | Cognitive function | Thinking abilities including ability to concentrate, remember, and to have awareness of others and the surrounding environment | Life impact | Cognitive Functioning |
| 4 | Comorbidity | Having more than one long term illness or condition occurring at the same time. Preventing comorbidity refers to preventing development of a new illness alongside the original health problem | Physiological or clinical | General Outcomes |
| 5 | Cost effectiveness** | The value for money of a treatment | Resource use | Economic |
| 6 | Cardiovascular risk | A person's future risk of developing problems with the heart or blood circulation, such as a heart attack or stroke | Physiological or clinical | Cardiac Outcomes |
| 7 | Cardiovascular event | Problems with the function of the heart or blood vessels such as a heart attack, stroke, abnormal heart rhythm, chest pain caused by reduced blood flow to the heart muscle, or heart failure | Physiological or clinical | Cardiac Outcomes |
| 8 | Diet | A person's regular eating habits | Physiological or clinical | Metabolism and Nutrition Outcomes |
| 9 | Exercise tolerance | How much exercise an individual can manage | Life impact | Physical Functioning |
| 10 | Fatigue | Extreme tiredness or lack of energy | Life impact | Physical Functioning |
| 11 | Health literacy | The ability to gather, read, understand, and use healthcare information to make informed decisions about one's own health | Life impact | Personal Circumstances |
| 12 | Healthcare use** | How much a person uses a healthcare service including visiting a doctor or other health professional or going to hospital | Life impact | Delivery of Care |
| 13 | Health-related quality of life | The impact of physical or mental health conditions and their treatments on how a person views their quality of life or wellbeing | Life impact | Perceived Health Status |
| 14 | Prevention of hypertension | Prevention of developing high blood pressure | Physiological or clinical | Vascular Outcomes |
| 15 | Psychological wellbeing | A person's emotional and mental wellbeing, including problems with low mood or anxiety | Life impact | Emotional Functioning/Wellbeing |
| 16 | Death | - | Death | Mortality/Survival |
| 17 | Organ damage | Some form of injury or harm leading to an organ such as lungs or kidneys not working well | Physiological or clinical | General Outcomes |
| 18 | Pain | - | Physiological or clinical | General Outcomes |
| 19 | Quality of life | A person's view of their emotional, physical, material, and social well-being. It includes the degree to which a person feels they are healthy, comfortable, and able to participate in or enjoy life events. | Life impact | Global Quality of Life |
| 20 | Reduced medication | Reducing the dose or number of drugs a person requires | Life impact | Delivery of Care |
| 21 | Timely screening | An activity that detects a health problem (or risk of a health problem) early, so it can be prevented or treated. | Resource use | Need for Further Intervention |
| 22 | Treatment satisfaction | How content a patient is with the treatment they have received, and how well they think it was delivered | Life impact | Delivery of Care |
| 23 | Weight | - | Physiological or clinical | Metabolism and Nutrition Outcomes |
| 24 | Obesity | A condition in which a person's weight is above a certain level, which is associated with risks to health, including increased risk of heart disease and diabetes | Physiological or clinical | Metabolism and Nutrition Outcomes |
| **Additional prevention outcomes added at Delphi R1** | | | | |
| 1 | Carer burden | Emotional, physical, and psychological stress or burden that caregivers experience when looking after a person with health problems. Caregivers may be family members, friends or health and social care workers | Resource use | Societal/Care Burden |
| 2 | Chronic disease self-management | The ability of an individual to undertake activities by themselves to manage their long-term health conditions | Life impact | Delivery of Care |
| 3 | Early detection | When a condition is identified early enough to prevent it from developing fully | Physiological or clinical | General Outcomes |
| 4 | Functioning/ADL | A person's ability to carry out activities of daily living for themselves e.g., washing or shopping | Life impact | Role Functioning; Physical Functioning; Social Functioning |
| 5 | Health anxiety | A fear or worry of becoming ill | Life impact | Perceived Health Status |
| 6 | Health risk behaviour | Behaviours that increase the risk of disease or injury e.g., smoking | Physiological or clinical | General Outcomes |
| 7 | Health seeking behaviour | Activities undertaken by an individual in order to receive treatment for a health problem | Life impact | Delivery of Care |
| 8 | Income | The money that a person receives on a regular basis in exchange for working or providing services | Life impact | Personal circumstances |
| 9 | Loneliness | A feeling of being alone caused by a lack of social contact | Life impact | Social Functioning |
| 10 | Perceived health | A person's own assessment of how good they feel their overall health is | Life impact | Perceived Health Status |
| 11 | Self-efficacy | An individual's belief to have the ability to do what is necessary to achieve a desired outcome (e.g., symptom control). | Life impact | Personal circumstances |
| 12 | Social functionality | How an individual fulfils their roles in and engages with their social environment (e.g., family, work) | Life impact | Social Functioning |
| **Treatment outcomes** | | | | |
| 1 | Acceptance of illness | Being able to come to terms with a diagnosis | Life impact | Emotional Functioning/Wellbeing |
| 2 | Adherence to treatment | The extent to which a person keeps to a recommended treatment, such as taking prescribed drugs or following a diet or exercise programme | Life impact | Delivery of Care |
| 3 | Adverse events | Any unexpected or unintended negative consequences of an intervention, for example, allergic reaction to a drug, or injury from a recommended exercise programme | Adverse events | Adverse Events/Effects |
| 4 | Aggression | A hostile behaviour towards another person that can lead to emotional or physical harm | Life impact | Emotional Functioning/Wellbeing |
| 5 | Agitation | Level of restlessness | Life impact | Emotional Functioning/Wellbeing |
| 6 | Appetite | - | Physiological or clinical | Metabolism and Nutrition Outcomes |
| 7 | Balance | A person's ability to stand or move without swaying or falling over | Physiological or clinical | Nervous System Outcomes |
| 8 | Carer burden*** | Emotional, physical, and psychological stress or burden that caregivers experience when looking after a person with health problems. Caregivers may be family members, friends or health and social care workers | Resource use | Societal/Care Burden |
| 9 | Cognitive function | Thinking abilities including ability to concentrate, remember, and to have awareness of others and the surrounding environment | Life impact | Cognitive Functioning |
| 10 | Comorbidity | Having more than one long term illness or condition occurring at the same time | Physiological or clinical | General Outcomes |
| 11 | Cost effectiveness | The value for money of a treatment | Resource use | Economic |
| 12 | Cardiac event risk | A person's future risk of developing problems with the heart or blood circulation, such as a heart attack or stroke | Physiological or clinical | Cardiac Outcomes |
| 13 | Cardiovascular event | Problems with the function of the heart or blood vessels such as a heart attack, stroke, abnormal heart rhythm, chest pain caused by reduced blood flow to the heart muscles, or heart failure | Physiological or clinical | Cardiac Outcomes |
| 14 | Diet | A person's regular eating habits | Physiological or clinical | Metabolism and Nutrition Outcomes |
| 15 | Domestic violence | Forms of abuse including physical and emotional, which occur within couple relationships or between family members in the home setting | Life impact | Personal circumstances |
| 16 | Emotional regulation | The ability to adjust one's emotions in response to a situation | Life impact | Emotional Functioning/Wellbeing |
| 17 | Falls risk | The risk of falling, based on assessing multiple risk factors, such as problems with thinking and memory, vision, balance, or mobility. Falls tend to occur more in older people. | Life impact | Physical Functioning |
| 18 | Fatigue | Extreme tiredness or lack of energy | Life impact | Physical Functioning |
| 19 | Functioning/ADL | A person's ability to carry out activities of daily living for themselves e.g., washing or shopping | Life impact | Role Functioning; Physical Functioning; Social Functioning |
| 20 | Health anxiety | A fear or worry of becoming ill | Life impact | Perceived Health Status |
| 21 | Health literacy | The ability to gather, read, understand, and use healthcare information to make informed decisions about one's own health | Life impact | Personal Circumstances |
| 22 | Health risk behaviour | Behaviours that increase the risk of disease or injury e.g., smoking | Physiological or clinical | General Outcomes |
| 23 | Healthcare cost | The cost of a healthcare service that is paid either by the government, the patient, or their family | Resource use | Economic |
| 24 | Healthcare access | Being able to access health services to improve one's health and wellbeing | Life impact | Delivery of Care |
| 25 | Healthcare quality | The standard of healthcare provided by a service | Life impact | Delivery of Care |
| 26 | Healthcare staff communication | Written and spoken communication by healthcare staff to patients and caregivers about the patient's health and healthcare | Life impact | Delivery of Care |
| 27 | Healthcare use | How much a person uses a healthcare service including visiting a doctor or other health professional or going to hospital | Life impact | Delivery of Care |
| 28 | Hospital admission | When an individual is admitted onto a hospital ward as an inpatient | Resource use | Hospital |
| 29 | Health-related quality of life | The impact of physical or mental health conditions on how a person views their quality of life or wellbeing | Life impact | Perceived Health Status |
| 30 | Hypertension | High blood pressure | Physiological or clinical | Vascular Outcomes |
| 31 | Illness resolution | When a health problem has fully resolved leading to that aspect of health returning to normal | Physiological or clinical | General Outcomes |
| 32 | Illness stigma | The negative attitudes people have towards someone with a particular physical or mental health condition | Life impact | Emotional Functioning/Wellbeing |
| 33 | Illness under control | A health problem that is no longer getting worse due to a treatment(s) | Physiological or clinical | General Outcomes |
| 34 | Income | The money that a person receives on a regular basis in exchange for working or providing services | Life impact | Personal circumstances |
| 35 | Increase in symptoms | The increase in physical or mental features of a health problem such as pain or shortness of breath | Physiological or clinical | General Outcomes |
| 36 | Loneliness | A feeling of being alone caused by a lack of social contact | Life impact | Social Functioning |
| 37 | Psychological wellbeing | A person's emotional and mental wellbeing, including problems with low mood or anxiety | Life impact | Emotional Functioning/Wellbeing |
| 38 | Death | - | Death | Mortality/Survival |
| 39 | Nausea | Feeling sick | Physiological or clinical | Gastrointestinal Outcomes |
| 40 | Pain | - | Physiological or clinical | General Outcomes |
| 41 | Perceived health | A person's own assessment of how good they feel their overall health is | Life impact | Perceived Health Status |
| 42 | Physical activity | Any body movement including sitting, standing, walking, running | Physiological or clinical | General Outcomes |
| 43 | Quality of life | A person's view of their emotional, physical, material, and social well-being. It includes the degree to which a person feels they are healthy, comfortable, and able to participate in or enjoy life events. | Life impact | Global Quality of Life |
| 44 | Reduced medication | Reducing the dose or number of drugs a person requires | Life impact | Delivery of Care |
| 45 | Self-management | The tasks that individuals must undertake themselves to manage long term health conditions | Life impact | Role Functioning |
| 46 | Sleep quality | How well someone sleeps | Physiological or clinical | General Outcomes |
| 47 | Treatment satisfaction | How content a patient is with the treatment they have received, and how well they think it was delivered | Life impact | Delivery of Care |
| 48 | Weight | - | Physiological or clinical | Metabolism and Nutrition Outcomes |
| 49 | Obesity | A condition in which a person's weight is above a certain level, which is associated with risks to health, including increased risk of heart disease and diabetes | Physiological or clinical | Metabolism and Nutrition Outcomes |
| **Additional treatment outcomes added at Delphi R1** | | | | |
| 1 | Continuity of care | - | Life impact | Delivery of Care |
| 2 | Frailty | - | Life impact | Social Functioning |
| 3 | Polypharmacy | The use of five or more routine medications - this can be over-the -counter, prescription, or complementary medicines | Physiological or clinical | General Outcomes |
| 4 | Self-esteem | An individual's own evaluation of their self-worth i.e., the confidence they have in themselves | Life impact | Emotional Functioning/Wellbeing |
| 5 | Social functionality | How an individual fulfils their roles in and engages with their social environment (e.g., family, work) | Life impact | Social Functioning |
| 6 | Treatment burden*** | The impact of the demands of treatment regimens on patient wellbeing, functioning, and time | Life impact | Personal circumstances |

* Dodd S, Clarke M, Becker L, Mavergames C, Fish R, Williamson PR. A taxonomy has been developed for outcomes in medical research to help improve knowledge discovery. Journal of clinical epidemiology. 2018;96:84-92.

** Under the prevention outcomes, there were discussions around the inclusion of ‘Cost effectiveness’ and ‘Healthcare use’, with prioritisation of the latter as it was considered a wider term that could include an estimate of the former. Although they did not make the final COS based on voting results, they were noted as important in some but not all studies and could be included as an additional outcome where appropriate.

*** Under the treatment outcomes, ‘Carer burden’ and ‘Treatment burden’ were individually excluded, but it was considered whether there should be a more generic outcome of ‘Burden’, with the narrative that the type of burden would be determined by the nature of the study. However, this was excluded by voting via email (62.5% votes).

### Appendix 6: Metrics/ measurement tools identified from the literature for the outcomes included in the two COS.

| **Outcomes** | **Metrics/ measurement tools used** |
| --- | --- |
| **Prevention of multimorbidity** | |
| Adverse events | Fatal (death, stroke), and non-fatal adverse events (hospitalisations, out-patient visits, surgeries etc.) |
| Comorbidity (development of new comorbidity) | Incidence of comorbidities |
| Health risk behaviour | Self-report measures of behaviour e.g., adherence to diet, exercise, medication etc.  Biochemical measures: Lipid profile, blood pressure, HbA1c etc.  Anthropometrics: height, weight, waist circumference etc. |
| Quality of life (including Health-related quality of life) | Short Form Health Survey (SF-36)  Short Form Health Survey (SF-12)  European Quality of Life (EQ-5D)  Health Utilities Index (HUI-3)  Saint George Respiratory Questionnaire hospital (SGRQ)  European Organization for Research and Treatment of Cancer Quality of Life Questionnaire (QLQ-C30) |
| **Treatment of multimorbidity** | |
| Adherence to treatment | Self-report measures such as Morisky Medication Adherence Scale (MMAS-4), CASE Adherence Index, Treatment Adherence Questionnaire of Patients with Hypertension (TAQPH), Visual Analogue Scale (VAS)  Other measures: Sputum collection with negative/positive sputum results, Wisepill (real-time electronic adherence monitoring) |
| Adverse events | Mortality rate  Treatment Emergent signs and symptoms (TESS) score  Incidence of adverse events (e.g., self-reported (e.g., vomiting), and/or biochemically ascertained (e.g., vital signs, electrocardiogram, haematological measures etc.) |
| Healthcare cost (out-of-pocket cost of treatment) | Direct non-medical (participant time spent travelling to and attending appointments), and indirect costs (lost productivity associated with illness or premature mortality). |
| Quality of life (including Health-related quality of life) | Generic questionnaires: World Health Organization Quality of Life- short version, WHOQOL-BREF, Short Form Health Survey (SF-12), Medical Outcomes Study 36-item, Short Form Health Survey (SF-36), European Quality of Life five-dimensional questionnaire, Five-level version (EQ-5D-5L).  Disease specific questionnaires: Irritable Bowel Syndrome Quality of Life (IBS-QOL) questionnaire, Hypertension scale of the Quality-of-Life Instruments for Chronic Diseases (QLICD-HY). |
